# Balancing donor health and plasma collection: a systematic review of the impact of plasmapheresis frequency

**DOI:** 10.1101/2024.03.05.24303709

**Authors:** Tine D’aes, Katja van den Hurk, Natalie Schroyens, Susan Mikkelsen, Pieter Severijns, Emmy De Buck, Peter O’Leary, Pierre Tiberghien, Veerle Compernolle, Christian Erikstrup, Hans Van Remoortel

## Abstract

Most plasma used for manufacturing plasma-derived medicinal products (PDMPs) such as albumin, immunoglobulin (Ig), and clotting factors is obtained from source plasma collected via plasmapheresis, the majority of which is contributed by the United States (US). While the demand for PDMPs continues to rise, it remains unclear whether high-frequency plasmapheresis, such as the twice-weekly plasma donation allowed in the US, may have any (long-term) adverse health effects on the donor.

To investigate the frequency at which plasma can be donated without harm to the donor, the current systematic review explores the impact of plasma donation frequency on cardiovascular health, protein depletion, and adverse events in healthy plasma donors. We asked the following research question: What is the impact of plasmapheresis frequency (Intervention) on the safety or health (Outcome) of healthy donors (Population)?

Six databases (PubMed, Embase, Web of Science, CINAHL, the Cochrane Library, and Transfusion Evidence Library), two clinical trial registries (ICTRP and clinicaltrials.gov), and the PROSPERO database were searched. Four observational and two experimental studies were included, and one ongoing randomized controlled trial was identified. The results showed that very high-frequency donation (twice per week) may result in a clinically relevant decrease in ferritin and bring IgG levels below the EDQM-defined lower threshold of 6 g/l. However, the evidence is of low to very low certainty, and solid conclusions are hindered by the *healthy donor effect* and methodological limitations of the included studies. In order to determine a safe threshold donation frequency that minimizes any possible harmful effect on the donor, more high-quality prospective cohort studies and experimental studies are thus needed. In the meantime, we argue for a precautionary approach and suggest that a sustainable and stable plasma supply may better rely on a large number of voluntary donors donating at a lower frequency (up to two donations per month), rather than on a small number of donors donating at a high frequency.

## Introduction

Plasma-derived medicinal products (PDMPs) such as albumin, immunoglobulin (Ig), and clotting factors have become essential in treating several disorders. The vast majority of plasma used for manufacturing PDMPs is obtained from source plasma that is collected through plasmapheresis, a procedure whereby plasma is removed while blood cells are returned to the donor. The United States (US) contributes around 70% of the global source plasma supply, which may be partly explained by its specific regulatory conditions, allowing for very high-frequency plasma donations, and its ability to remunerate plasma donors [1]. The recent COVID-19 crisis and concurrent decrease in donors highlighted the risks of Europe’s dependency on the US for plasma supplies, particularly as the demand for PDMPs such as IgG continues to rise [2, 3]. To address this issue, the European Blood Alliance (EBA) initiated the “*Strengthening voluntary non-remunerated plasma collection capacity in Europe*” (SUPPLY) project [4]. The main objective of this project is to develop recommendations and guidance for blood establishments, competent authorities, medical societies, and other professional stakeholders to support them in increasing plasma collection in the European Union (EU) while guaranteeing the safety of patients as well as donors.

Gaining European independence in the supply of PDMPs and upscaling plasma donation in Europe can logically be achieved by recruiting additional donors and/or by increasing donation frequency among existing donors. However, ensuring the safety of plasma donors in this process is of fundamental importance. Therefore, one of the work packages of the SUPPLY project focuses on the development of evidence-based recommendations for plasma donor protection practices. To this end, we have previously published a scoping review and evidence gap map (https://cebap.org/storage/cebap/schroyens-2023-egm.html), providing an overview of the available evidence and evidence gaps concerning plasma donor safety, plasma donation-related adverse or health effects, and plasma donor protection practices [5].

Although plasmapheresis is generally well-tolerated, potential long-term health consequences such as lasting effects on donor plasma protein levels and bone mineral density require further investigation [6]. Specifically, it remains unclear whether high-frequency plasmapheresis may pose any harm to donors. While plasma donors in the US can donate up to twice a week, allowing for a maximum of 104 donations each year, many European countries such as France, the Czech Republic, Italy, and the Netherlands limit plasmapheresis to once every two weeks, although Germany allows up to 60 donations per year [7, 8]. Claims that donating plasma twice a week, as allowed by the Food and Drug Administration (FDA), is proven to be safe [9] have often been based on insufficient evidence, mostly involving methodologically flawed observational studies that do not, in fact, support such conclusions [10].

To investigate the frequency at which plasma can be donated without causing harm to the donor, the current systematic literature review analyses, synthesizes, and critically appraises the best available evidence regarding the effect of plasmapheresis frequency on donor safety. Secondary outcomes of interest include cardiovascular health and protein levels.

## Methods

This review was registered prospectively in the International Prospective Register of Systematic Reviews (PROSPERO; https://www.crd.york.ac.uk/prospero/) with ID 405419 (CRD42023405419). Any amendments to the protocol are described and explained under the heading ‘deviations from the initial protocol’. In addition, the review was conducted in accordance with the methodological charter of the Centre for Evidence-Based Practice (CEBaP) [11]. The Preferred Reporting Items for Systematic Reviews and Meta-Analyses (PRISMA) checklist was used to guide reporting.

Our research question, structured according to the PICO framework (Patient-Intervention-Comparator-Outcome), is: What is the impact of plasmapheresis frequency (I) on the safety or health (O) of healthy donors (P)?

### Eligibility criteria

#### Publication type and study design

Articles published in peer-reviewed journals and clinical trial registrations were considered eligible for inclusion. On the other hand, conference abstracts or papers, dissertations not accompanied by a peer-reviewed publication, editorials, and letters to the editor were excluded. Peer-reviewed publications or clinical trial registrations reporting on study protocols for which no results were available (yet) were not included but labelled as ‘awaiting classification’. These may be reconsidered in any future updates of this review.

The following study designs were eligible for inclusion: controlled experimental studies (i.e. randomized or non-randomized controlled trials) and controlled observational studies (i.e. controlled before-after studies, controlled interrupted time series, case-control studies, and cohort studies).

Narrative reviews were not included as such, but their reference lists were screened for relevant studies. Uncontrolled/descriptive studies (e.g. uncontrolled before-after studies, uncontrolled interrupted time series, case reports, case series), animal studies, and *ex vivo* or *in vitro* studies were excluded.

#### Population

Studies involving healthy (i.e. eligible to donate plasma) adults, both remunerated and non-remunerated, who donated plasma via plasmapheresis (consisting of the withdrawal of blood, separation of plasma from blood cells, and return of blood cells to the body) were eligible for inclusion. Studies with multicomponent apheresis (plasma, platelets, and red blood cells, or any combinations of these), a mixed donor population (whole blood, plasma, and/or platelets) with no separate data for plasma donors, and patient populations who underwent autologous plasmapheresis or therapeutic plasma exchange and/or received PDMPs were excluded.

#### Intervention-comparator

We included studies comparing donors undergoing plasmapheresis at a higher frequency (shorter intervals between sessions) to donors undergoing less frequent or no plasmapheresis. A clear quantitative description of the frequency (i.e. number of donations per time unit) of plasma donation in the intervention and comparator groups had to be available (e.g. weekly donation versus donation every two weeks).

Studies with whole blood or platelet donation as the only comparator, studies that provide no quantitative information on the frequency of plasma donation (e.g. studies comparing first-time versus repeat donors), and studies that compare different (cumulative) numbers of donations (i.e. without specifying the exact donation frequency, even if an average donation frequency can be estimated based on the cumulative number of donations over a certain period), were excluded.

#### Outcomes

The primary outcome of interest was the occurrence of adverse events, including all events classified in the 2014 Standard for Surveillance of Complications Related to Blood Donation developed by hemovigilance experts from the American Association of Blood Banks (AABB), the International Society of Blood Transfusion (ISBT), and the International Hemovigilance Network (IHN) [12, 13]. In addition, other (long-term) adverse events such as lowered bone density/osteoporosis, vein fibrosis (inability for venepuncture) or skin fibrosis were included. For more details, we refer to the protocol registered in PROSPERO (CRD42023405419) [14].

Secondary outcomes included cardiovascular health (encompassing both biochemical risk markers and physiological or clinical risk factors) and protein levels (including any protein measured in blood, serum or plasma).

Quantitative data on the outcomes had to be present in the paper in order for it to be included in the current systematic review.

#### Other criteria

There were no restrictions regarding the date of publication. Language was limited to publications in English, Dutch, French or German, i.e. languages that could be unequivocally interpreted by the reviewers.

### Search strategy

A scoping review [5] preceded the current systematic review. For this scoping review, an initial search for eligible studies was performed on 10 October 2022. For the current systematic review, these search strategies were re-run on 4 December 2023 with a publication date filter starting from 10 August 2022 to ensure an overlap of two months with the search conducted for the scoping review. The following resources were searched from the time of inception up to 4 December 2023: MEDLINE, PMC, and NCBI bookshelf (via the PubMed interface), Embase (via the Embase.com interface), Web of Science Core Collection (Science Citation Index Expanded (SCI-EXPANDED) and Conference Proceedings Citation Index-Science (CPCI-S)), CINAHL (via the EBSCO interface), Cochrane Library (systematic reviews and controlled trials), Transfusion Evidence Library, International Clinical Trials Registry Platform (ICTRP), Clinicaltrials.gov, and PROSPERO.

Search strings, consisting of combinations of indexing terms (MeSH terms for Medline, Cochrane CENTRAL and CINAHL, Emtree terms for Embase) and free text words, related to the population and outcomes of the research question, were developed per database and are shown in Appendix A. It should be noted that, as mentioned, these search strings were developed for the initial scoping review, and as such covered a broader research question than the topic addressed in this systematic review.

For all studies included in this systematic review, we screened the reference lists and the 20 first ‘similar articles’ in PubMed for additional relevant records.

### Study selection

Records retrieved from the database/trial registry searches were uploaded to EndNote [15]. Duplicate records were removed using the automatic deduplication function of EndNote, followed by a manual check for accuracy and additional duplicates. All records were independently screened for eligibility by two reviewers (HVR, TD, NS, and/or PS) at the level of the title and abstract, followed by full-text screening. Disagreements were resolved by discussion, and where necessary by consulting a third reviewer (TD or HVR) or our panel with subject matter expertise (KVDH, CE, VC, PT, PO).

### Data extraction

Data from included studies were extracted independently by two reviewers (HVR and TD), using an *a priori*-developed data extraction form (Appendix B). Disagreements in extracted data were resolved by discussion, where necessary by consulting a third reviewer (NS), to eventually result in a completed consensus data extraction form. The following information was extracted from the selected studies:

- Study characteristics: author, year, country, study sponsor, financial disclosures of the co-authors, study design, and descriptions of the population, intervention and comparator
- Study findings: outcomes, comparison (intervention versus comparator), effect sizes, and the number of participants

Continuous outcome data were expressed as means ± standard deviations (unless otherwise reported). Dichotomous outcome data were expressed as risk ratios (RRs) with 95% confidence intervals (CIs) (unless otherwise reported). Results were considered statistically significant if p<0.05. Effects were considered clinically relevant if values fell outside the normal range or were determined to be significant by our expert panel. If results were available for different time points, data from all time points were extracted.

For extraction of the data from the study by Mortier et al. [16], NS replaced HVR since HVR was one of the co-authors of the study. TD and NS were not involved in this study.

Where possible, the authors of the included studies were contacted via e-mail to obtain missing information or data.

### Risk of bias and Grading of Recommendations, Assessment, Development and Evaluation (GRADE) assessment

For each study, the risk of bias was assessed by two reviewers independently (HVR and TD, or, for the study by Mortier et al. [16], NS and TD) using the Grading of Recommendations, Assessment, Development and Evaluation (GRADE) key criteria (Appendix C) [17]. Limitations that may result in bias according to the GRADE approach include ‘inappropriate eligibility criteria’, ‘inappropriate methods for exposure variables’, ‘not controlling for confounding’, ‘incomplete or inadequate follow-up’, and ‘other limitations’ in case of observational studies. For experimental studies, these include ’lack of randomization’ and ‘lack of allocation concealment’, ‘lack of blinding’, ‘incomplete accounting of outcome events’, ‘selective outcome reporting’, and ‘other limitations’ [18]. Discrepancies between reviewers were resolved through discussion.

Next, the GRADE approach was used to assess the overall certainty of the body of evidence as ‘high’, ‘moderate’, ‘low’ or ‘very low’. Experimental studies receive an initial grade of ‘high’ and can be downgraded based on the risk of bias, imprecision, inconsistency, indirectness, and publication (i.e. non-reporting) bias. Observational studies receive an initial grade of ‘low’ and can be downgraded based on the risk of bias, imprecision, inconsistency, indirectness, and publication (i.e. non-reporting) bias, or upgraded based on large effect, dose–response gradient, and plausible confounding [19].

### Data synthesis

Meta-analyses could not be conducted since no studies were sufficiently comparable in intervention and outcomes. Statistical synthesis of these results was deemed inappropriate, and no statements about the consistency of effects across studies or outcomes were made to avoid unintentional vote counting [20]. Summarizing conclusions were formulated according to the certainty of the evidence, which is reflected in the wording of the statements [21, 22].

### Deviations from the initial protocol

The PICO question specified in the protocol has been adapted from “Does frequent plasmapheresis (I) affect the safety or health (O) of plasma donors (P)?” to “What is the impact of plasmapheresis frequency (I) on the safety or health (O) of healthy donors (P)?” to clarify the fact that only studies comparing different, specified donation frequencies or comparing a specified donation frequency to a no-donation control group were eligible for inclusion. The intention of the review and inclusion criteria have remained unchanged.

Contrary to what was written in the protocol, we did not include studies comparing first-time plasmapheresis donors with repeat donors since we decided that this comparison would not allow us to investigate the effect of donation frequency.

## Results

### Search results

For the initial scoping review [5], 17810 records were identified from databases and registers. After removal of duplicates, 7209 records were screened, resulting in the inclusion of 97 papers and registrations. An additional five records were identified through reference lists of narrative reviews. Among the 102 records included in the scoping review (94 research articles, one study protocol and seven registrations), five articles and one ongoing experimental study were eligible for inclusion in the current systematic review. The updated search identified an additional 2124 hits (1351 after the removal of duplicates), leading to the inclusion of one additional study. Published studies that met the inclusion criteria described above, were withheld for data extraction. A total of six studies were thus included in the review and one ongoing experimental study was labelled as ’awaiting classification’. Figure 1 shows the detailed flow diagrams with the number of records at each step of the study selection process for both the initial scoping review and the current systematic review.

**Figure 1:**
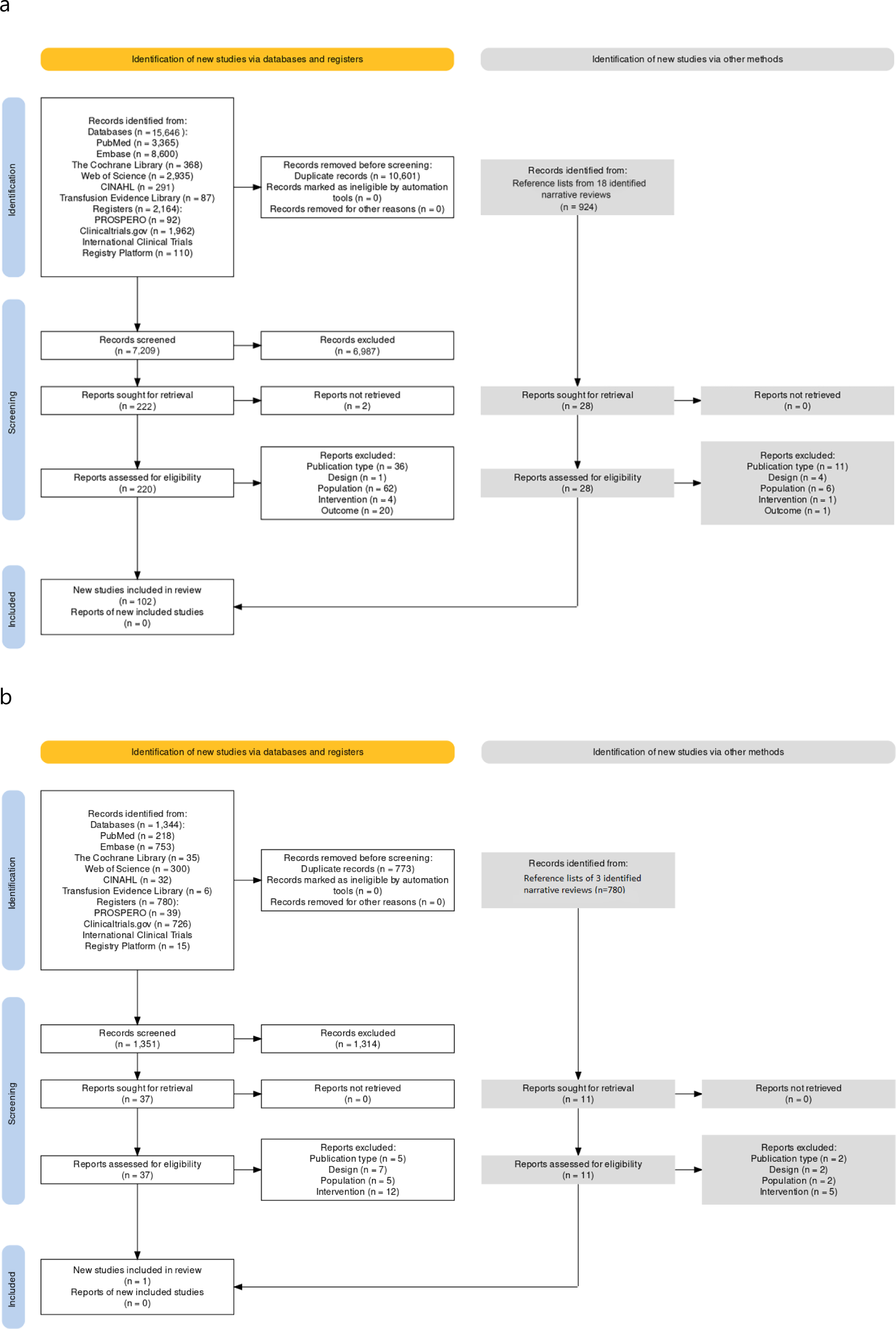
PRISMA study selection flow diagrams for the scoping review (a) and update for the systematic review (b).

### Characteristics of included studies

Of the six included studies, two studies were conducted in Croatia [23, 24], two in the US [25, 26], one in Canada [27], and one in Belgium [16]. The studies comprised four observational [23–26], and two experimental studies [16, 27]. In addition, we found one registration of a randomized controlled trial (RCT) that will be conducted in Norway [28] (Table 1). The included studies were mostly older, with one study conducted in the early 1970s [26], two in the early 1980s [23, 24], one in the early 1990s [27], one in the early 2010s [25] and only one very recent paper [16]. For an overview of the study characteristics, see Table 1.

**Table 1:**
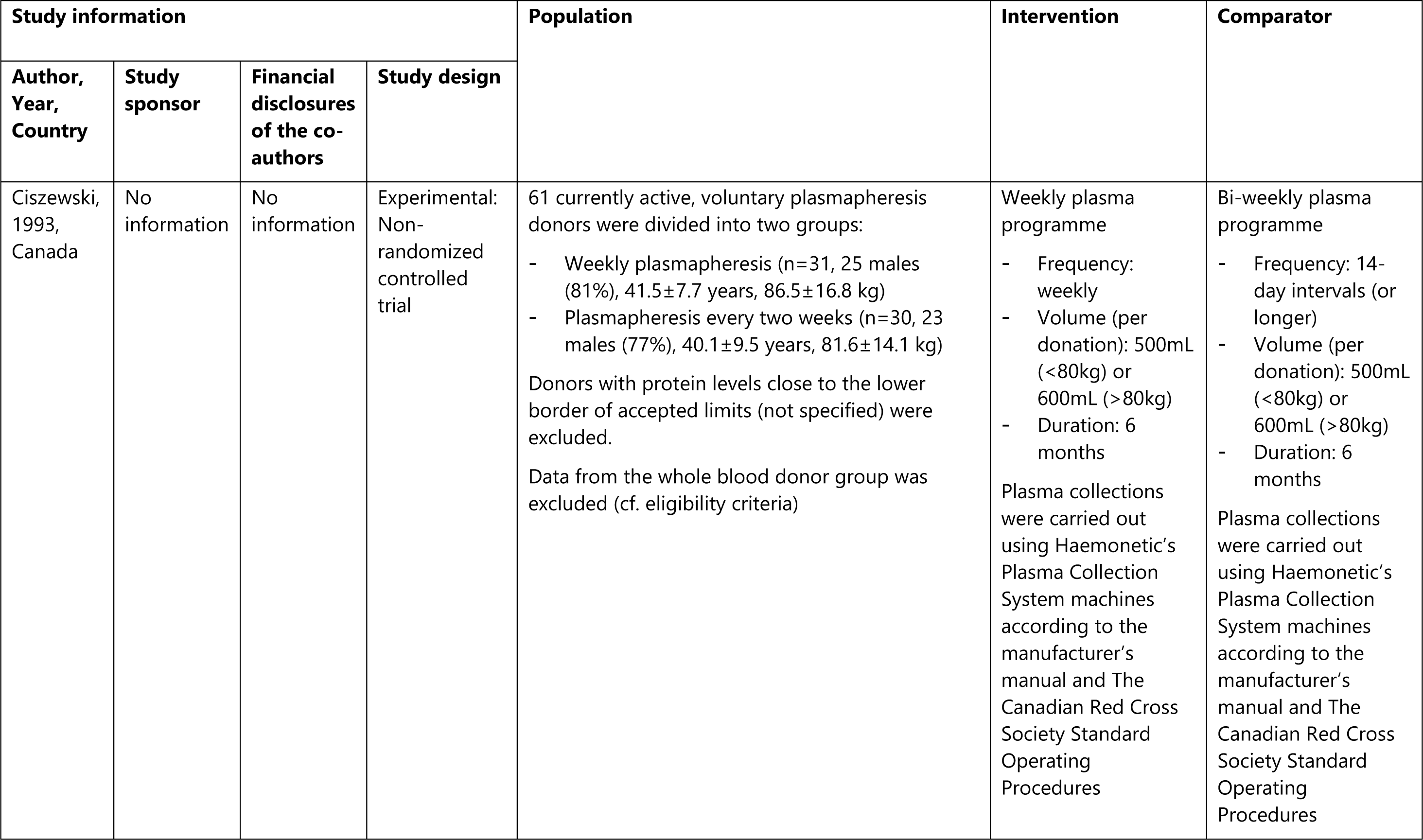
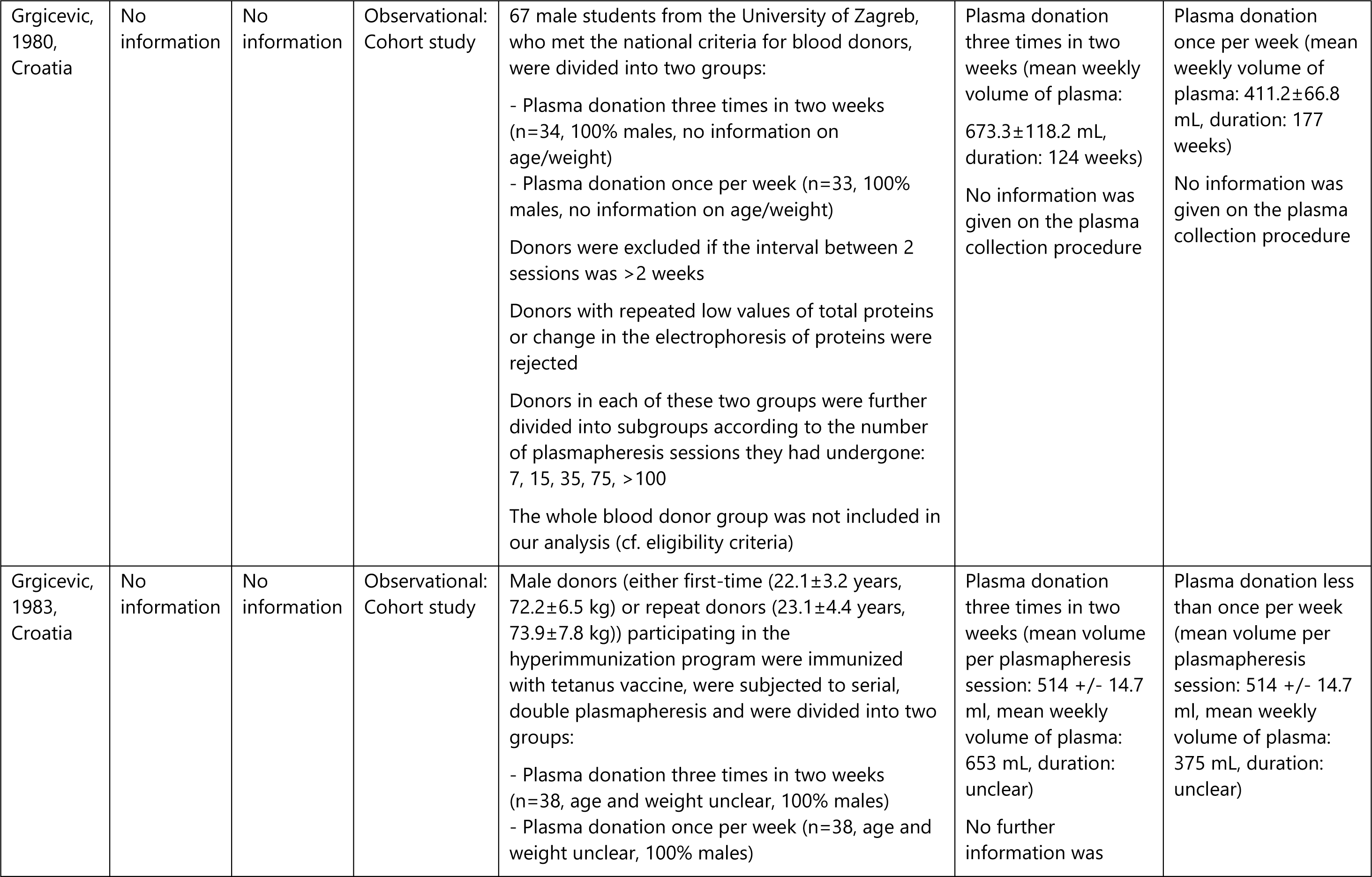
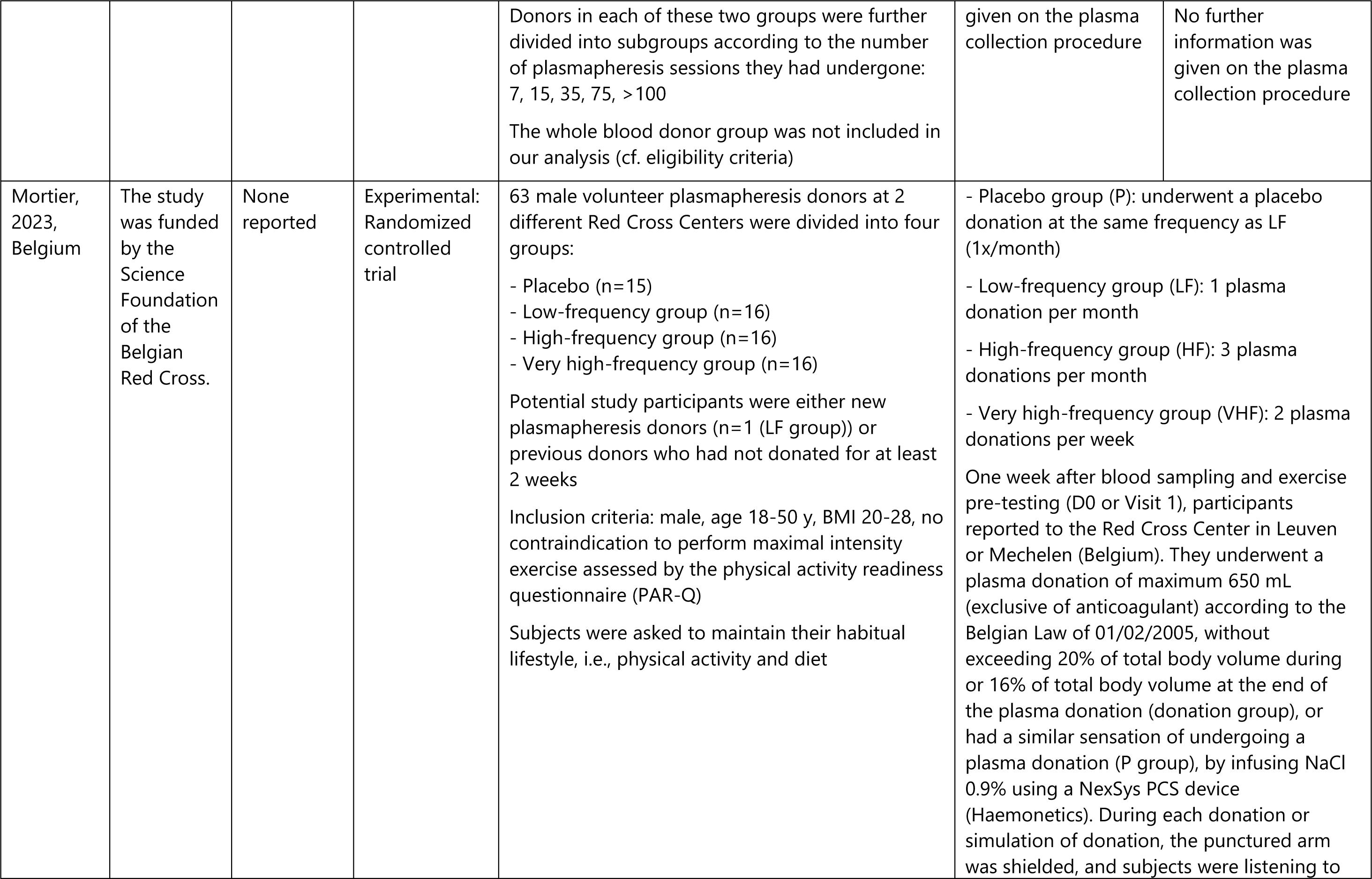
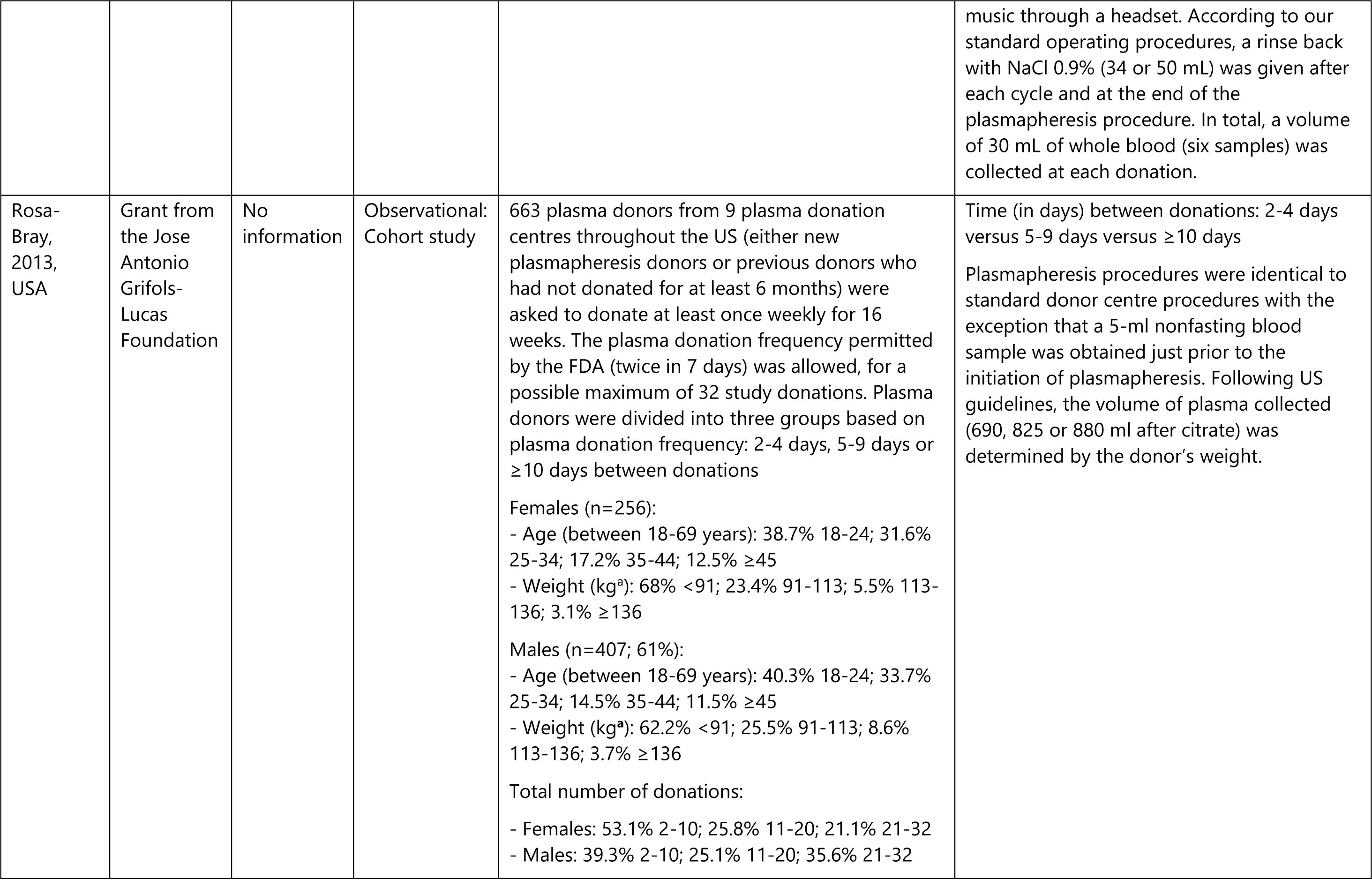
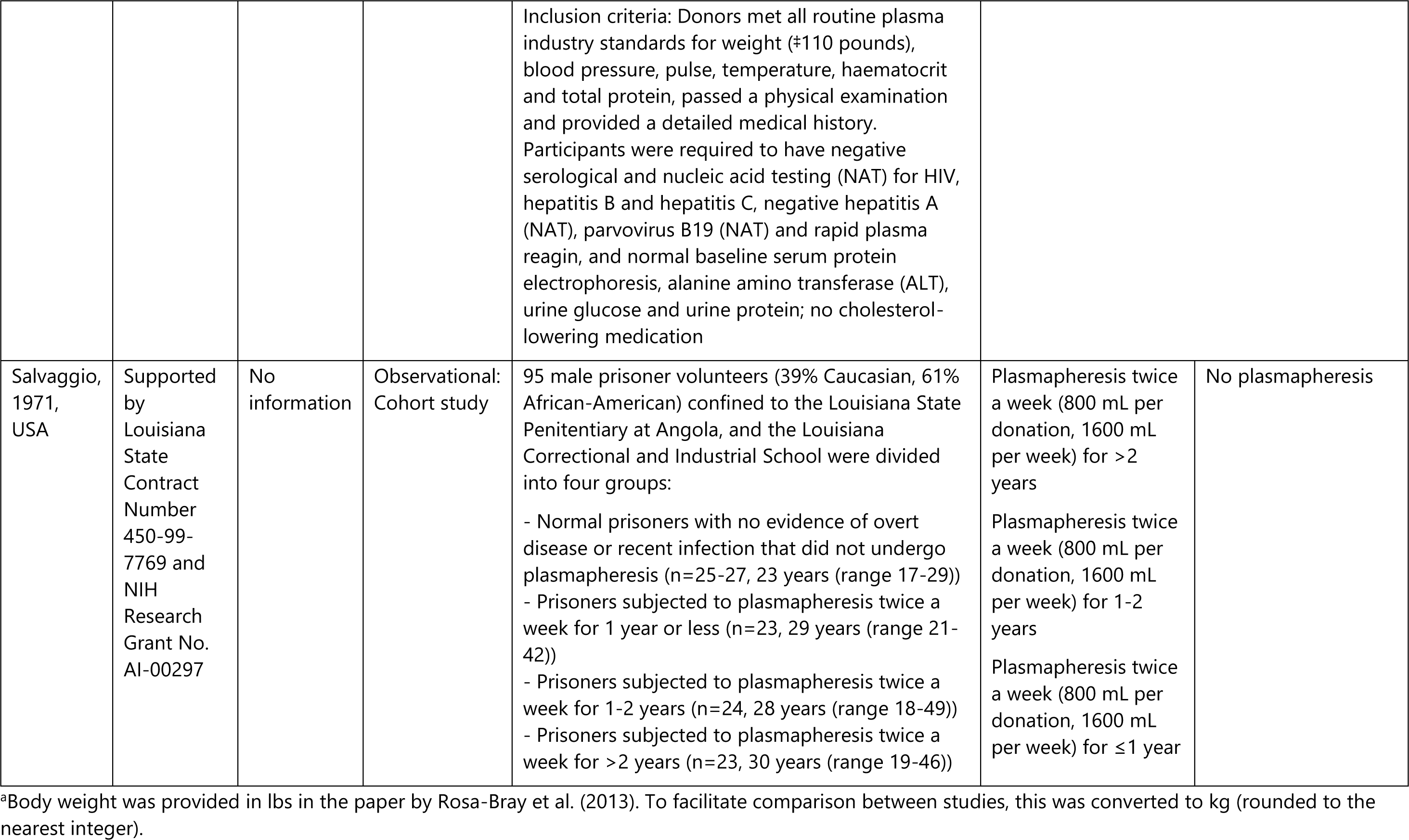
Characteristics of included studies.

For the two experimental studies, the donor population consisted of currently active plasma donors (except for one first-time donor in the study by Mortier et al.) [16, 27]. One of the observational studies involved either new donors or donors who had not donated for at least six months [25], one included students with unspecified donation histories [24], one involved prisoners that had been donating for less than one your, for one to two years, or for more than two years [26], and one study involved both first-time and repeat donors [23]. Four of the six studies exclusively included male donors, one study included 61% males [25], and the sixth study had a population that was 77-81% male [27].

Among the four included observational studies, there were two studies originating from the same research group that compared donors undergoing plasmapheresis at a frequency of three times per two weeks with weekly (or less frequent) plasmapheresis [23, 24]. One study compared three different intervals between subsequent plasma donations over a period of 16 weeks: two to four days, five to nine days, and ten days or more [25]. One study compared the effects of two plasma donations a week for one year or less, for one to two years, or for more than two years with no plasmapheresis [26]. The two experimental studies comprised one study comparing weekly plasmapheresis with plasmapheresis every 14 days for six months [27], and one study comparing four different conditions: a monthly sham plasmapheresis procedure, monthly plasmapheresis, plasmapheresis three times a month and plasmapheresis twice a week for 12 weeks [16].

The Norwegian RCT registration is set to assess the effects of plasmapheresis three times per two weeks versus every two weeks (versus whole blood donation every three months) during 16 weeks in male donors [28].

Only one study reported on the occurrence of adverse events, our primary outcome, in groups of plasma donors donating at different frequencies. The authors distinguished haematoma, vasovagal reactions, anaemia (defined as a haemoglobin level below 135 g/l), and ’other’ adverse events [16].

The following outcomes relating to cardiovascular health were assessed in the included studies: change in low-density lipoprotein (LDL), high-density lipoprotein (HDL) or total cholesterol [25], glycemia [16], insulinemia [16], glycated haemoglobin (HbA1C) [16], total cholesterol [16], systolic blood pressure [16], diastolic blood pressure [16], body mass [16], body mass index (BMI) [16], bone mineral content [16], fat mass [16], fat-free mass [16], fat-free mass + bone mineral content [16], fat percentage [16], lactate during cycling at 190 W [16], lactate after cycling [16], maximum heart rate [16], maximal ventilation [16], peak oxygen consumption [16], and oxygen pulse [16].

Levels of the following proteins were measured: total serum protein [24, 27], IgG [16, 24, 26, 27], IgA [16, 24, 26, 27], IgM [16, 24, 26, 27], albumin [16, 24, 26], (alpha 1 or 2, beta and gamma) globulins [23, 24, 26], alpha-1-antitrypsin [23, 24], alpha-2-macroglobulin [23, 24], anti-thrombin III [23, 24], plasminogen [23, 24], fibrinogen [23, 24], factor V [23], transferrin [26], ceruloplasmin [26], haptoglobin [26], factor VIII [23], alkaline phosphatase [24], glutamic oxaloacetic transaminase (GOT) [24], glutamic-pyruvic transaminase (GPT) [24], haemoglobin [16], ferritin [16], C-reactive protein (CRP) [16], and creatine kinase [16].

The Norwegian RCT registration is set to investigate total protein, IgG, other plasma proteins, and inflammation markers [28].

### Risk of bias of included studies and GRADE assessment

#### Experimental studies

The two experimental studies both had very small sample sizes. The study by Mortier et al. [16] performed a sample size calculation for the outcome total serum protein only and used a sample size that corresponded to this calculated optimal information size.

Both studies suffered from several limitations in study design (Figure 2 & Appendix C). The study by Ciszewski et al. [27] did not randomly allocate donors to the intervention groups. Although participants in the study by Mortier et al. [16] did not know whether they were in the intervention or sham group, they did of course know the frequency at which they came to the centre. Neither study mentioned blinding of personnel and outcome assessors. Both experimental studies suffered from limited compliance with the plasma donor regimen. Dropout rates differed across groups for both studies and neither study included the data of dropouts in their statistical analyses. The study by Ciszewski et al. [27] failed to provide detailed information on differences in adverse events and albumin levels between groups. Another limitation in both studies is that all (but one) donors were known donors, which may result in the selection of a population better able to cope with potential side effects of plasmapheresis.

**Figure 2:**
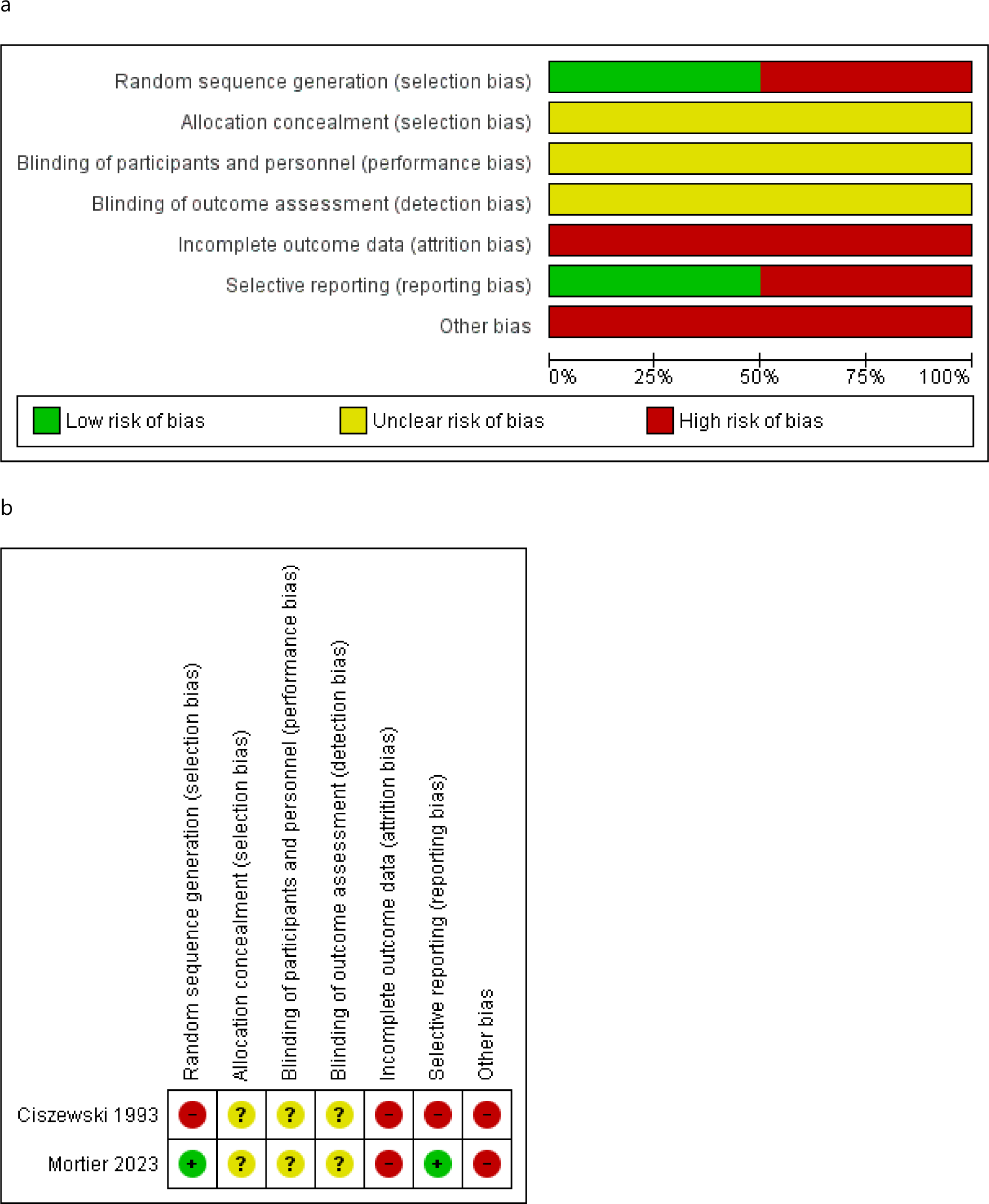
Review authors’ judgments on the risk of bias for the two included experimental studies, based on the GRADE criteria. Risk of bias is presented per item, either as percentages across all included studies (a) or separately for each included study (b).

Overall, the body of evidence for the experimental studies was downgraded by one for limitations in study design. In addition, evidence was downgraded by one for imprecision, given the limited sample sizes. Results were not downgraded for indirectness, even if the generalizability of the studies, comprising active, relatively young and (predominantly) male donors, is unsure. This resulted in the classification of the certainty of evidence as “low” for the experimental studies (Appendix C).

#### Observational studies

All included observational studies had small sample sizes and none performed a sample size calculation. The observational studies lacked controlling for confounding (except for one [25]), suffered from high dropout rates (except one study [26], but this might have been a retrospective study), failed to mention clear inclusion criteria [23, 24, 26], neglected donors with low protein levels [24], did not mention the donation history of donors [23, 24], or did not specify when outcomes were assessed exactly [24, 26].

The observational evidence was thus downgraded by one for study limitations (Figure 3) and by one for imprecision due to limited sample sizes. The evidence was not downgraded for indirectness, even if external validity may have been limited because the papers were mostly old, and predominantly used young male students or prisoners. This resulted in the classification of the certainty of evidence as “very low” for the observational studies (Appendix C).

**Figure 3:**
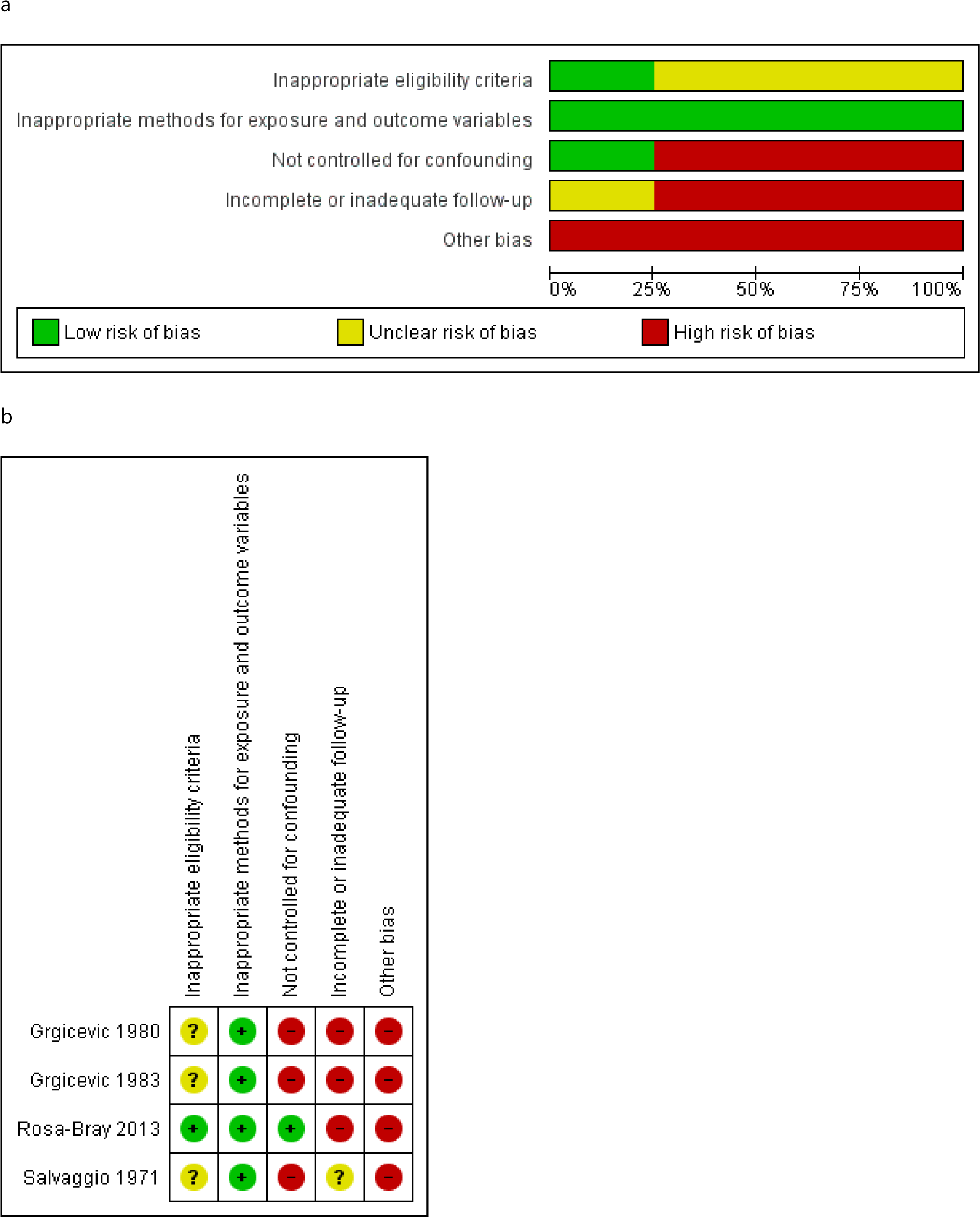
Review authors’ judgments on the risk of bias for the four included observational studies, based on the GRADE criteria. Risk of bias is presented per item, either as percentages across all included studies (a) or separately for each included study (b).

### Synthesis of results

Results were split according to the plasma donation frequencies under study into five different comparisons: (1) plasmapheresis twice per week versus three times per month versus once per month versus placebo [16], (2) weekly plasmapheresis versus plasmapheresis every two weeks [27], (3) three plasma donations per two weeks versus (less than) one per week [24, 29], (4) plasmapheresis twice per week versus no plasmapheresis for more than two years, one to two years, or one year or less [26], and (5) plasmapheresis intervals of 2-4 days versus 5-9 days versus 10 days or more [25]. Detailed summaries of the results for the five individual comparisons can be found in Appendix D.

## Plasmapheresis twice per week (n=16) versus three times per month (n=16) versus once per month (n=16) versus placebo (n=15): one study, 63 participants (Mortier, 2023)

### Primary outcomes

#### Adverse events

The RCT by Mortier et al. [16] was the only included study to investigate the effect of plasmapheresis frequency on adverse event occurrence. No adverse events were recorded in donors undergoing sham or monthly plasmapheresis. Five haematomas in total were present in donors who donated three times a month (three events in three donors, adverse event rate: 1.08/50 donations) or two times a week (two events in one donor, adverse event rate: 0.28/50 donations). A total of five vasovagal reactions were reported: one in a donor who donated three times a month (adverse event rate: 0.36/50 donations) and four in three donors who donated two times a week (adverse event rate: 0.57/50 donations). Five anaemia events, defined as a haemoglobin level below 135 g/l, were detected in four of the donors who donated twice a week (adverse event rate: 0.71/50 donations). No other (major) events were reported.

### Secondary outcomes

#### Cardiovascular health

No statistically significant interaction effect between the donation frequency group and the timepoint at which the outcomes were measured (one week before the first donation, 42 days after the first donation, or 84 days after the first donation) was found for the following outcomes related to cardiovascular health: total cholesterol (p=0.39), insulinemia (p=0.40), systolic (p=0.15) and diastolic (p=0.90) blood pressure, body mass (p=0.37), BMI (p=0.33), bone mineral content (p=0.08), fat mass (p=0.70), fat-free mass (p=0.76), fat-free mass + bone mineral content (p=0.79), fat percentage (p=0.75), lactate at 190W (p=0.92), lactate post-exercise (p=0.88), maximal heart rate (p=0.94), maximal ventilation (p=0.86), peak oxygen consumption (p=0.79), and oxygen pulse (p=0.54) [16]. Differences that were not statistically significant, were also not clinically relevant.

On the other hand, a statistically significant group*time interaction effect was found for glycemia (p=0.036) and glycated haemoglobin (HbA1C; p=0.0007). For glycemia, significant differences were found for three plasma donations per month (78.6±14.8 mg/dl) compared to one donation per month (92.4±22.4 mg/dl; p=0.02) or to placebo (91.4±17.82 mg/dl; p=0.03), and for two plasma donations per week (79.9±14.4 mg/dl) compared to one donation per month (92.4±22.4 mg/dl; p=0.03) or to placebo (91.4±17.82 mg/dl; p=0.04). For HbA1C, a significant effect was found only between donors who made two donations per week (5.1±0.4%) and those who made one donation per month (5.3±0.4%; p=0.03). While statistically significant, these effects on HbA1C and on glycemia were not considered to be clinically relevant [16].

#### Protein levels

No significant group*time interaction effect was found for C-reactive protein (p=0.95) and creatine kinase (p=0.45). A statistically significant group*time interaction effect was found for albumin (p<0.0001), haemoglobin (p<0.0001), ferritin (p=0.0014), IgA (p<0.0001), IgG (p<0.0001), and IgM (p<0.0001). For IgA and IgM, despite a significant group*time interaction effect, there was no statistical significance for any of the pairwise comparisons between the different frequency groups [16].

For albumin, a significant reduction was found with two plasma donations per week (40.6±2.4 g/l) in comparison with either three donations per month (44.7±3.2 g/l; p<0.0001), one donation per month (43.7±2.8 g/l; p=0.001) or with placebo (45.9±2.7 g/l; p<0.0001). In addition, albumin levels were significantly lower in participants donating once a month (43.7±2.8 g/l) than in participants who underwent a sham donation (45.9±2.7 g/l; p=0.03). However, none of the effects on albumin levels were considered to be clinically relevant.

Donating twice a week significantly reduced haemoglobin levels (13.9±0.8 g/dl) compared to donating three times a month (15.0±0.8 g/dl; p=0.001), once a month (14.5±0.8 g/dl; p=0.046), or via a sham procedure (15.1±0.77 g/dl; p=0.0001). Again, the effects on haemoglobin levels were not considered to be of clinical significance.

Donating three times a month (31.0±14.8 µg/l) or twice a week (20.1±10 µg/l) both resulted in a large decrease in ferritin levels compared to donating once a month (74.6±66 µg/l; respective p-values: p=0.01 and p=0.001) or to a sham procedure (82.7±55.77 µg/l; respective p-values: p=0.003 and p=0.0003). Of note, a rinse back with 0.9% NaCl was given after each cycle and at the end of the donation procedure, and six samples of 30 ml of whole blood were taken at each donation.

Donating plasma twice a week (5.73±1.4 g/l) resulted in a large reduction in IgG levels compared to donating plasma three times a month (8.43±1.52 g/l; p<0.0001), once a month (8.7±2.2 g/l; p<0.0001) or compared to a sham procedure (10.59±1.86 g/l; p<0.0001), bringing IgG levels beneath the lower limit of normal of 6 g/l [30]. In addition, IgG was significantly reduced in participants who donated plasma three times (8.43±1.52 g/l) or once (8.7±2.2 g/l) per month compared to those who underwent a sham procedure (10.59±1.86 g/l; respective p-values: p=0.001 and p=0.005). There was no significant difference in IgG levels between participants donating three times a month and those donating on a monthly basis (p=0.68) [16].

### Summarized evidence conclusions

Very high-frequency plasmapheresis (up to twice a week) may result in little to no difference (either no effect or a small, unimportant effect) in the levels of albumin, haemoglobin, IgA, IgM, C-reactive protein, and creatine kinase compared to less frequent or sham apheresis. While effects on albumin and haemoglobin levels may be statistically significant, these were not considered to be clinically relevant [16].

Plasmapheresis three times a month or two times a week may result in a large reduction in ferritin levels compared to less frequent or sham plasmapheresis. Donating twice a week may even bring ferritin levels below the threshold for iron deficiency, which (in women) may lie at 15 µg/l or 25 µg/l depending on the source [31].

Plasmapheresis twice a week may result in a large reduction of IgG to levels below the lower limit of 6 g/l [30], compared to less frequent or sham plasmapheresis [16].

For factors related to cardiovascular health, it was found that highly frequent plasmapheresis (up to twice a week) may result in little to no difference (either no effect or a small, unimportant effect) in insulinemia, HbA1C, glycemia, total cholesterol, systolic blood pressure, diastolic blood pressure fat percentage, lactate at 190W, lactate post-exercise, maximal heart rate, maximal ventilation, peak oxygen consumption, and oxygen pulse, body mass, BMI, bone mineral content, fat mass, fat-free mass, and fat-free mass + bone mineral content [16].

From the available evidence, a difference in the occurrence of adverse events between different frequencies of plasmapheresis could not be demonstrated.

The evidence is of low certainty and cannot be considered precise due to limited sample sizes.

## Weekly plasmapheresis (n=31) versus plasmapheresis every two weeks (n=30): one study, 61 participants (Ciszewski, 1993)

### Secondary outcomes

#### Protein levels

Donating weekly did not affect levels of IgG (p=0.12-0.17), IgA (p=0.13-0.70) or IgM (p=0.05-0.53) in comparison with donating every two weeks. A small decrease in total serum protein was observed in participants donating weekly compared to participants donating once every two weeks at three (64.6±2.6 g/l vs 67.9±3.7 g/l; p=0.004) and at six months (66.2±3.0 g/l vs 69.5±3.0 g/l; p=0.002) only, but reduces values were within the normal ranges and were therefore not considered to be clinically relevant. No significant differences were observed in total serum protein levels after one (p=0.84), two (p=0.69), four (p=0.11) or five (p=0.27) months of plasma donation [27].

### Summarized evidence conclusions

Donating plasma once a week may result in little to no difference in total serum protein, IgG, IgA, or IgM levels compared to donating once every two weeks [27]. The evidence is of low certainty and cannot be considered precise due to limited sample sizes.

## Plasmapheresis three times every two weeks versus (less than) once per week: two studies, total number of participants unclear (Grgicevic, 1980&1983)

### Secondary outcomes

#### Protein levels

No significant differences were observed for total serum protein (p=0.82), IgG (p=0.45), IgA (p=0.90), IgM (p=0.65), albumin (p=0.88-0.94), alpha-1&2, beta(−1), or gamma globulins (p=0.20-0.93), alpha-1-antitrypsin (p=0.16-0.97), alpha-2-macroglobulin (p=0.09-0.98), anti-thrombin III (p=0.07-0.98), fibrinogen (p=0.26-1.00), factor VIII (p=0.13-0.96), alkaline phosphatase (p=0.19), GOT (p=0.92), or GPT (p=0.96) between participants donating three times per two weeks in comparison with participants who donated once per week (or less) [23, 24]. For plasminogen, a significant difference was observed between both groups after seven donations (79.10±22.62% vs 96.56±12.60%; p=0.01), but not after one (p=0.97), 15 (p=0.63), 35 (p=0.73), 75 (p=0.22) or more than 100 donations (p=0.47) [23]. For factor V, significant differences were observed after 7 (0.87±0.07 vs 0.93±0.07 u/l x 10^3^; p=0.04), 15 (0.80±0.01 vs 0.88±0.11 u/l x 10^3^; p=0.03) and 75 donations (0.91±0.11 vs 0.84±0.11 u/l x 10^3^; p=0.03). After seven and 15 sessions, levels were lower for donors donating three times every two weeks compared with donors donating (less than) once a week, but after 75 donations the opposite was observed [23]. These differences were not considered to be clinically meaningful.

### Summarized evidence conclusions

The evidence is very uncertain about the effect of plasmapheresis three times every two weeks compared to weekly plasmapheresis on the levels of total protein, IgG, IgA, IgM, albumin, globulins (alpha-1&2, beta(−1), gamma), alpha-1-antitrypsin, alpha-2-macroglobulin, anti-thrombin III, plasminogen, fibrinogen, factor V, factor VIII, alkaline phosphatase, GOT, and GPT [23, 24]. The evidence is of very low certainty and results cannot be considered precise due to limited sample sizes.

## Plasmapheresis twice per week versus no plasmapheresis (n=27) for more than two years (n=23), one to two years (n=24), or one year or less (n=23): one study, 97 participants (Salvaggio, 1971)

### Secondary outcomes

#### Protein levels

No effects were observed on the levels of ceruloplasmin (p=0.35-1.00) and haptoglobin (p=0.13-0.54). Levels of IgG, IgA, IgM, albumin and B1-globulin were significantly lower in plasma donors donating twice a week for either less than one year, between one and two years, or for more than two years than in non-donors.

For IgG, average levels were 8.60±1.83 g/l for donors donating for more than two years (p<0.00001), 7.32±1.87 g/l for donors donating for one to two years (p<0.00001), and 7.67±1.55 for donors donating for less than one year (p<0.00001), compared to 11.80±2.92 g/l in non-donors. In addition, donors who had been donating for over two years had slightly lower IgG levels than donors who had been donating for one to two years (p=0.02), but this difference was not considered to be clinically meaningful [26]. Furthermore, the number of participants with IgG levels below the lower limits of the normal range (determined in comparison with groups without plasmapheresis (normal Louisiana prisoners) and Behring values (healthy European civilians)) was significantly higher in donors donating for more than two years (11/23; p=0.01), for one to two years (16/24; p=0.002), or for less than one year (15/23; p=0.003) than in non-donors (2/25).

For IgA, average levels were 1.32±0.27 g/l for donors donating for more than two years (p=0.01), 1.20±0.32 g/l for donors donating for one to two years (p=0.002), and 1.28±0.43 g/l for donors donating for less than one year (p=0.01), compared to 1.78±0.89 g/l in non-donors. In addition, the number of participants with IgA levels below the lower limits of the normal range was significantly higher in donors donating for one to two years (14/24; p=0.01), or for less than one year (13/23; p=0.02), than in non-donors (5/25). There was no statistically significant difference in the number of participants with low IgA levels between donors donating for more than two years (10/23) and non-donors (5/25; p=0.10).

For IgM, average levels were 0.75±0.33 g/l for donors donating for more than two years (p=0.0007), 0.88±0.46 g/l for donors donating for one to two years (p=0.03), and 0.71±0.18 g/l for donors donating for less than one year (p<0.0001), compared to 1.20±0.59 g/l in non-donors. In addition, the number of participants with IgM levels below the lower limits of the normal range was significantly higher in donors donating for more than two years (17/23; p=0.003), for one to two years (14/24; p=0.02), or for less than one year (17/23; p=0.003), than in non-donors (6/25).

For albumin, average levels were 26.17±2.32 g/l for donors donating for more than two years (p<0.00001), 25.74±3.80 g/l for donors donating for one to two years (p<0.00001), and 27.73±3.23 for donors donating for less than one year (p=0.0001), compared to 31.99±4.57g/l in non-donors.

For B1-globulin, average levels were 0.65±0.16 mg% for donors donating for more than two years (p<0.0001), 0.64±0.13 mg% for donors donating for one to two years (p<0.00001), and 0.70±0.14 mg% for donors donating for less than one year (p=0.002), compared to 0.83±0.16 mg% in non-donors.

For transferrin, levels were significantly decreased only in donors donating for less than one year (170±21 mg/dl) compared to non-donors (193±46 mg/dl; p=0.02), but not in donors donating for one to two (p=0.13) or more than two years (p=0.09) [26].

### Summarized evidence conclusions

The evidence is very uncertain about the effect of donating plasma twice a week compared to not donating plasma on the levels of transferrin, ceruloplasmin, and haptoglobin. Donating plasma twice a week may reduce levels of IgG, IgA, IgM, albumin, and B1-globulin compared to not donating plasma, but the effects may not be clinically meaningful and the evidence is very uncertain. Donating plasma twice a week may increase the number of donors with IgG, IgA, and IgM levels below the lower limit of normal, but the evidence is very uncertain [26]. Results cannot be considered precise due to limited sample sizes.

## Interval between plasma donations of 2-4 days versus 5-9 days versus 10 days or more: one study, 663 participants (Rosa-Bray, 2013)

### Primary outcomes

#### Adverse events

The study by Rosa-Bray only reported on the total number of adverse events (14, including eight mild and six moderate) across all participants across the three different frequency groups, but did not provide separate data for different frequency groups [25]. These data were therefore not extracted.

### Secondary outcomes

#### Cardiovascular health

Total cholesterol was lower (larger decrease or smaller increase) for participants donating at a frequency of 2-4 days between donations (between −46.6±5.2 and −5±1.1) compared to those donating every 5-9 days (between −32±5 and +6.9±1.5; p≤0.00001-0.0002) or ≥ 10 days (between − 20.8±5.1 and +7.8±1; p<0.00001 for all subgroups), and for 5-9 days compared to ≥ 10 days (p≤0.00001-0.0004) between donations, for both males and females with high (≥240), higher than desired (200-239), or acceptable (<200) baseline cholesterol levels. Significance was only not reached for the comparison between 5-9 days and ≥10 days in males with high baseline cholesterol levels (p=0.12).

LDL cholesterol was lower for participants donating at a frequency of 2-4 days between donations (between −35.3±5 and −1.9±0.6) compared to those donating every 5-9 days (between −20.8±4.8 and +5.4±1; p≤0.00001-0.0001) or ≥ 10 days (between −25.6±5 and +6.8±1.2; p≤0.00001-0.004), and for 5-9 days compared to ≥ 10 days between donations (p≤0.00001-0.0006), for both males and females with high (≥160), higher than desired (130-159), or acceptable (<130) baseline LDL cholesterol levels. Significance was only not reached for the comparison between 5-9 days and ≥10 days in males (p=0.53) and females (p=0.11) with high baseline LDL cholesterol levels.

HDL cholesterol was lower for participants donating at a frequency of 2-4 days between donations (between −10.8±1.5 and 0±0.7) compared to those donating every 5-9 days (between −6.7±1.5 and +4.7±0.8; p≤0.00001-0.01) or ≥ 10 days (between −4±1.5 and +4.9±0.5; p<0.00001 for all comparisons), and for 5-9 days compared to ≥ 10 days between donations (p≤0.00001-0.0008), for both males and females with low (<40), average (4-60 for males, 5-60 for females), or optimal (>60) HDL baseline cholesterol levels. Significance was only not reached for the comparison between 5-9 days and ≥10 days in males with low (p=0.25) or average (p=1.00) baseline HDL cholesterol levels [25].

### Summarized evidence conclusions

Donating every 2-4 days, compared to donating every 5-9 days or every 10 days or less, or donating every 5-9 days compared to donating every 10 days or less, may reduce levels of total, LDL and HDL cholesterol [25]. The evidence is of very low certainty and cannot be considered precise due to limited sample sizes.

## Discussion

This paper is the first systematic review to study the effect of plasmapheresis frequency on donor health and safety, including appraisal of the certainty of all available evidence using the internationally recognized GRADE approach. We retrieved six eligible studies, including two experimental and four observational studies. We found limited evidence indicating that three plasmapheresis procedures per month or more may result in a clinically relevant reduction in ferritin. Twice-weekly plasmapheresis may even bring ferritin levels below the threshold for iron deficiency, which (in women) may lie at 15 µg/l or 25 µg/l depending on the source [31]. In addition, plasmapheresis twice a week may result in a clinically relevant reduction of IgG to levels below the lower threshold of normal of 6 g/l [30]. No (clinically relevant) effects could be demonstrated for any of the other outcomes. Although the primary outcome of interest of this review was the occurrence of adverse events, only one study with very small sample sizes provided information on adverse event rates in different donation frequency groups [16]. Since all evidence was of low to very low certainty, further research is very likely to have an important impact on the effect estimates.

The observation that twice-weekly plasmapheresis may bring IgG levels below 6 g/l stands out among the results. Previous cohort studies [32, 33] have also indicated a potentially significant depletion of IgG with high-frequency plasmapheresis. The authors of the *Study on Intensive Plasmapheresis* (SIPLA) conducted in Germany concluded that long-term intensive donor plasmapheresis is safe, although 12.4% of experienced donors were excluded due to low (<5.8 g/l) IgG levels [33]. A follow-up study, including both first-time and experienced donors, showed that IgG levels fell below the 6 g/l limit in 27.1% of donors who made an average of 29.8 (±24.8) donations (over an unclear period of time), again not preventing the authors from concluding that the investigated plasmapheresis frequency was safe [34]. Similarly, although not necessarily related to IgG levels, the authors of a recent study concluded that high-frequency plasmapheresis is safe, despite the observation that 45.5% of ceased donors reported health issues or concerns potentially related to plasmapheresis, arguing that it was not *the* most often reported reason to cease donating [35]. The health consequences of hypo-IgG in plasma donors remain to be elucidated (e.g. [36, 37]) and is unclear whether (acute) hypo-IgG leads to an increased infection risk in this population [6]. Future prospective cohort studies and randomized controlled trials should be initiated to examine the health consequences of hypo-IgG levels. In the meantime, we argue for a precautionary principle, prioritizing donor safety until more scientific evidence is available [38].

A limitation of the evidence body is that four of the six included studies were observational studies of very low certainty, which are subject to the so-called *healthy donor effect*, i.e. the observation that donors are healthier than the average population since they have to meet strict eligibility criteria [39]. Subsequently, through multiple donations, there is a selection of donors who can withstand frequent donations. The higher the donation frequency, the stronger the healthy donor effect may be. This phenomenon may conceal deleterious effects, especially if donors who cease donating are not followed up. To mitigate this bias, RCTs and studies taking donors’ pre-existing health status into account are recommended [39].

In addition, generalization of the results is further limited by the lack of results in women. Four of the six included studies, in addition to the ongoing RCT, had an entirely male donor population, and another included study had a population that was 77-81% male [27]. The only study that included 39% females [25], exclusively investigated cholesterol levels.

Notably, this review identified only two published experimental studies, including just one RCT, as well as one ongoing Norwegian RCT assessing the effects of different plasmapheresis frequencies [28]. To assess long-term effects, well-conducted prospective cohort studies providing real-world data should ideally be combined with long RCTs. Studies such as that performed by Di Angelantonio et al. (2007) on whole blood donors [40] show that large RCTs are feasible, although keeping donors committed to donating plasma twice a week for a prolonged period of time may be highly challenging. To facilitate the set-up of these studies and ensure comparable big data sets across different countries, we also recommend obliging the implementation of a register for standardized haemovigilance data within Europe [38].

Strict monitoring and deferral of donors with, for example, low IgG levels may play an important role in preventing harmful effects for the donor. On the other hand, existing deferral practices may also result in the underestimation of the side effects of plasmapheresis. The European Directorate for the Quality of Medicines & HealthCare (EDQM) recommends measuring IgG every 26 donations and at least yearly [30], but this may not be frequent enough to pick up on fast-occurring drops in IgG levels. Furthermore, the EDQM suggests using tests to assess iron status, such as ferritin, to prevent iron depletion in donors [30]. In addition to screening active donors, it is important to follow up on donors after they are deferred for low protein levels, to study recovery and the likelihood of their return as donors. Examining dropout rates and reasons and following up on lost donors will help to better understand the impact of frequent plasmapheresis. The follow-up period in the study by Mortier [16] was limited to three months, so it is unclear how protein levels evolved afterward. A lot of value may lie in long-term monitoring of donor health through databases and health registers (cf. [41]). Future research should generate evidence to inform optimal IgG algorithms and test intervals, which is currently lacking [38]. In addition, in frequent donors, it may be important to limit the loss of red blood cells, and consequently of iron and ferritin, by rinsing back at the end of the procedure and limiting whole blood sampling for test purposes to a minimum [16, 30].

We did not include studies that compared the health of first-time or new donors to that of repeat donors [42, 43] or investigated health effects in donors with different cumulative numbers of plasma donations [44, 45]. We did, however, identify these studies systematically in the framework of our scoping review [5]. Several retrospective studies found no effect of the number of donations made in the previous 12 months on iron levels, protein levels, immunity markers, red cell and iron metabolism, cardiovascular risk, or risk of (osteoporotic) fractures [41, 46, 47]. One of these studies did find that donors who made an average of 16 to 55 donations (depending on the group) in the previous 12 months had significantly lower total serum protein, albumin, and IgG levels than non-donors [47]. Such retrospective studies are subject to several limitations, including the healthy donor effect and missing data. Even if the number of donations over a certain period reported in some of these studies may allow calculating an average donation frequency, it is impossible to differentiate, for example, donors who donated at a very high frequency for only a short period of time from donors who donated at a low frequency for the whole study period based on this information. Although these studies may also contain relevant information for refining the regulatory framework surrounding plasmapheresis, they did not directly address the current research question and were thus not included in this review. The studies that were included explicitly aimed to compare different donation frequencies, despite potential imperfect adherence to the donation regimens.

In conclusion, although the precise effects of high-frequency plasmapheresis on donor health remain to be elucidated, the current evidence shows that it may significantly affect donor IgG and ferritin levels. In addition, it is important to underscore the lack of robust evidence supporting the safety of such frequent plasma donation. Pending additional high-quality prospective and experimental studies, a sustainable increase in plasma donation may better rely on a large number of voluntary donors donating at a lower frequency than on a low number of donors donating at a high frequency. Based on the limited scientific evidence available today, we recommend a maximum of two plasma donations per month, pending sufficient evidence confirming the safety of higher donation frequencies. This recommendation stems from the precautionary principle, prioritizing donor safety until more information is available [38].

## Data Availability

The authors confirm that the data supporting the findings of this study are available within the article and its supplementary materials.

## Acknowledgements

Funding: this work was funded by the EU4Health Programme of the European Union (“project 101056988/SUPPLY”). The content of this manuscript represents the views of the authors only and is their sole responsibility; it cannot be considered to reflect the views of the European Commission and/or the European Health and Digital Executive Agency (HaDEA) or any other body of the European Union. The European Commission and the Agency do not accept any responsibility for use that may be made of the information it contains.

In addition, this work was co-funded by the Foundation for Scientific Research of the Belgian Red Cross.

We acknowledge the following members of SUPPLY Work Package 5 for providing input throughout the different steps in conducting this systematic review: Thomas Burkhardt (German Red Cross), Elodie Pouchol (Etablissement Français du Sang), Pascale Richard (Etablissement Français du Sang), Marloes Spekman (Sanquin), Petar Kos (European Blood Alliance), and Torsten Tonn (German Red Cross).

## References

[1] Hartmann J, Klein HG. Supply and demand for plasma-derived medicinal products - A critical reassessment amid the COVID-19 pandemic, Transfusion 2020; 60:2748–52. https://10.1111/trf.16078.

[2] Brand A, De Angelis V, Vuk T, Garraud O, Lozano M, Politis D. Review of indications for immunoglobulin (IG) use: Narrowing the gap between supply and demand, Transfus Clin Biol 2021; 28:96–122. https://10.1016/j.tracli.2020.12.005.

[3] Domanovic D, von Bonsdorff L, Tiberghien P, Strengers P, Hotchko M, O’Leary P, Thibert JB, Magnussen K, Erikstrup C, Spekman M, Chesneau S, Jones J, Moller BK, Verheggen P, Gogarty G, Elzaabi M, de Angelis V, Candura F, Mali P, Rossi F, Rodrigues B, Sepetiene R, Lenzen T, Walsemann S, Perry R, Plançon JP, So-Osman C, Durand-Zaleski I, Facco G, Thijssen-Timmer D. Plasma collection and supply in Europe: Proceedings of an International Plasma and Fractionation Association and European Blood Alliance symposium, Vox Sang 2023; 118:798–806. https://10.1111/vox.13491.

[4] European Blood Alliance Strengthening plasma collection in Europe. 2022. https://europeanbloodalliance.eu/strengthening-plasma-collection-in-europe/ (Accessed: 15/09/2022

[5] Schroyens N, D’Aes T, De Buck E, Mikkelsen S, Tiberghien P, van den Hurk K, Erikstrup C, Compernolle V, Van Remoortel H. Safety and protection of plasma donors: A scoping review and evidence gap map, Vox Sang 2023. https://10.1111/vox.13544.

[6] Hoad VC, Castrén J, Norda R, Pink J. A donor safety evidence literature review of the short-and long-term effects of plasmapheresis, Vox Sang 2023. https://10.1111/vox.13512.

[7] Purohit M, Berger M, Malhotra R, Simon T. Review and assessment of the donor safety among plasma donors, Transfusion 2023; 63:1230–40. https://10.1111/trf.17369.

[8] Strengers PFW. Challenges for Plasma-Derived Medicinal Products, Transfus Med Hemother 2023; 50:116–22. https://10.1159/000528959.

[9] Plasma Protein Therapeutics Association (PPTA). Study confirms that frequency of source plasma donation as regulated by U.S. FDA does not impair donor health and well-being. 2023. https://www.pptaglobal.org/material/study-confirms-that-frequency-of-source-plasma-donation-as-regulated-by-u-s-fda-does-not-impair-donor-health-and-well-being#:∼:text=Study%20Confirms%20that%20Frequency%20of,Donor%20Health%20and%20Well%2D Being (Accessed: 07/12/2023

[10] Van Remoortel H, van den Hurk K, Compernolle V, O’Leary P, Tiberghien P, Erikstrup C. Very-high frequency plasmapheresis and donor health-absence of evidence is not equal to evidence of absence, Transfusion 2023; 63:2358–61. https://10.1111/trf.17601.

[11] CEBaP. Development of evidence-based guidelines and systematic reviews: methodological charter. 2020. https://www.cebap.org/storage/cebap/inf-methodology-charter-cebap.pdf. (Accessed: February 1 2023).

[12] International society of blood transfusion working party on haemovigilance network and the AABB donor haemovigilance working group. Standard for surveillance of complications related to blood donation. 2014. https://www.aabb.org/docs/default-source/default-document-library/resources/donor-standard-definitions.pdf?sfvrsn=21834fa4_0 (Accessed: 9 February 2023

[13] Townsend M, Kamel H, Van Buren N, Wiersum-Osselton J, Rosa-Bray M, Gottschall J, Rajbhandary S. Development and validation of donor adverse reaction severity grading tool: enhancing objective grade assignment to donor adverse events, Transfusion 2020; 60:1231–42. https://10.1111/trf.15830.

[14] Van Remoortel H, D’aes T, Schroyens N, De Buck E, van den Hurk K, Erikstrup C, Compernolle V, Mikkelsen S, Tiberghien P, O’Leary P, Torsten T, Pouchol E, Spekman M, Richard P, Burkhardt T. The impact of frequent plasmapheresis on adverse events, cardiovascular health, and protein levels in plasma donors: a systematic review of controlled experimental and observational studies. 2023. https://www.crd.york.ac.uk/prospero/display_record.php?RecordID=405419 (Accessed: 2023 Jul 25

[15] Lee D, Thornton P, Hirst A, Kutikova L, Deuson R, Brereton N. Cost effectiveness of romiplostim for the treatment of chronic immune thrombocytopenia in Ireland, Applied health economics and health policy 2013; 11:457–69. https://10.1007/s40258-013-0044-y.

[16] Mortier A, Khoudary J, van Dooslaer de Ten Ryen S, Lannoy C, Benoit N, Antoine N, Copine S, Van Remoortel H, Vandekerckhove P, Compernolle V, Deldicque L. Effects of plasmapheresis frequency on health status and exercise performance in men: A randomized controlled trial, Vox Sang 2023. https://10.1111/vox.13569.

[17] Schünemann H, Brożek J, Guyatt G, Oxman A. Handbook for grading the quality of evidence and the strength of recommendations using the GRADE approach. Updated October 2013., Place Publishe.

[18] Guyatt GH, Oxman AD, Vist G, Kunz R, Brozek J, Alonso-Coello P, Montori V, Akl EA, Djulbegovic B, Falck-Ytter Y, Norris SL, Williams JW, Jr., Atkins D, Meerpohl J, Schunemann HJ. GRADE guidelines: 4. Rating the quality of evidence--study limitations (risk of bias), J Clin Epidemiol 2011; 64:407–15. https://10.1016/j.jclinepi.2010.07.017.

[19] Atkins D, Best D, Briss PA, Eccles M, Falck-Ytter Y, Flottorp S, Guyatt GH, Harbour RT, Haugh MC, Henry D, Hill S, Jaeschke R, Leng G, Liberati A, Magrini N, Mason J, Middleton P, Mrukowicz J, O’Connell D, Oxman AD, Phillips B, Schunemann HJ, Edejer T, Varonen H, Vist GE, Williams JW, Jr., Zaza S, Group GW. Grading quality of evidence and strength of recommendations, BMJ 2004; 328:1490. https://10.1136/bmj.328.7454.1490.

[20] McKenzie J, Brennan S. Chapter 12: Synthesizing and presenting findings using other methods. In: T.J. Higgins JPT, Chandler J, Cumpston M, Li T, Page MJ, Welch VA editor. Cochrane Handbook for Systematic Reviews of Interventions. Version 6,0.: Cochrane; 2019. Available from www.training.cochrane.org/handbook.

[21] Santesso N, Glenton C, Dahm P, Garner P, Akl EA, Alper B, Brignardello-Petersen R, Carrasco-Labra A, De Beer H, Hultcrantz M, Kuijpers T, Meerpohl J, Morgan R, Mustafa R, Skoetz N, Sultan S, Wiysonge C, Guyatt G, Schunemann HJ, Group GW. GRADE guidelines 26: informative statements to communicate the findings of systematic reviews of interventions, J Clin Epidemiol 2020; 119:126–35. https://10.1016/j.jclinepi.2019.10.014.

[22] Schünemann HJ, Vist GE, Higgins JPT, Santesso N, Deeks JJ, Glaziou P, Akl EA, Guyatt GH. Chapter 15: Interpreting results and drawing conclusions. In: J.P.T. Higgins, J. Thomas, J. Chandler, M. Cumpston, T. Li, M.J. Page, V.A. Welch editors. Cochrane Handbook for Systematic Reviews of Interventions version 6.0 (updated July 2019). Cochrane: 2019.

[23] Grgicević D. Influence of long-term plasmapheresis on blood coagulation, Ric Clin Lab 1983; 13:21–31.

[24] Grgicević D, Pistotnik M, Pende B. Observation of the changes of plasma proteins after long term plasmapheresis, Dev Biol Stand 1980; 48:279–86.

[25] Rosa-Bray M, Wisdom C, Wada S, Johnson BR, Grifols-Roura V, Grifols-Lucas V. Prospective multicentre study of the effect of voluntary plasmapheresis on plasma cholesterol levels in donors, Vox Sang 2013; 105:108–15. https://10.1111/vox.12031.

[26] Salvaggio J, Arquembourg P, Bickers J, Bice D. The effect of prolonged plasmapheresis on immunoglobulins, other serum proteins, delayed hypersensitivity and phytohemagglutinin-induced lymphocyte transformation, Int Arch Allergy Appl Immunol 1971; 41:883–94. https://10.1159/000230580.

[27] Ciszewski TS, Ralston S, Acteson D, Wasi S, Strong SJ. Protein levels and plasmapheresis intensity, Transfus Med 1993; 3:59-65. https://10.1111/j.1365-3148.1993.tb00105.x.

[28] Sykehuset Innlandet HF. The Effect of Donation Frequency on Donor Health in Blood Donors Donating Plasma by Plasmapheresis. 2023.

[29] Grgicevic D. Influence of long-term plasmapheresis on blood coagulation, Ric Clin Lab 1983; 13:21–31.

[30] European Directorate for the Quality of Medicines & HealthCare (EDQM). Guide to the preparation, use and quality assurance of blood components. Recommendation No. R (95) 15. 21st edition. 2023.

[31] Addo OY, Mei Z, Hod EA, Jefferds ME, Sharma AJ, Flores-Ayala RC, Spitalnik SL, Brittenham GM. Physiologically based serum ferritin thresholds for iron deficiency in women of reproductive age who are blood donors, Blood Adv 2022; 6:3661–5. https://10.1182/bloodadvances.2022007066.

[32] Laub R, Baurin S, Timmerman D, Branckaert T, Strengers P. Specific protein content of pools of plasma for fractionation from different sources: impact of frequency of donations, Vox Sang 2010; 99:220–31. https://10.1111/j.1423-0410.2010.01345.x.

[33] Schulzki T, Seidel K, Storch H, Karges H, Kiessig S, Schneider S, Taborski U, Wolter K, Steppat D, Behm E, Zeisner M, Hellstern P, group Ss. A prospective multicentre study on the safety of long-term intensive plasmapheresis in donors (SIPLA), Vox Sang 2006; 91:162–73. https://10.1111/j.1423-0410.2006.00794.x.

[34] Kiessig ST, Teichmann S, Schneider S, Ouarrak T, Krause KP, Storch H, Hellstern P. First results from the Study on Intensive Plasmapheresis II (SIPLA II). 23rd Regional Congress of the ISBT. Amsterdam, The Netherlands: 2013.

[35] Fransen M, Becker M, Hershman J, Lenart J, Simon TL. Why do US source plasma donors stop donating?, Transfusion 2023; 63:1904–15. https://10.1111/trf.17522.

[36] Moog R, Laitinen T, Taborski U. Safety of Plasmapheresis in Donors with Low IgG Levels: Results of a Prospective, Controlled Multicentre Study, Transfusion Medicine and Hemotherapy 2022. https://10.1159/000522528.

[37] Preußel K, Offergeld R. Comment to Moog et al.: Safety of Plasmapheresis in Donors with Low IgG Levels: Results of a Prospective, Controlled Multicentre Study, Transfus Med Hemother 2022; 49:404–5. https://10.1159/000526175.

[38] Erikstrup C, Van Remoortel H, van den Hurk K, Schroyens N, D’aes T, Spekman M, Mikkelsen S, Compernolle V, Tonn T, Burkhardt T, Gubbe K, Tiberghien P, Pouchol E, Richard P, O’Leary P. Deliverable 5.3: Recommendations on protection of plasma donors. 2023. https://supply-project.eu/wp-content/uploads/2024/02/D5.3-Recommendations-on-protection-of-plasma-donors.pdf (Accessed: 2024 February 28

[39] van den Hurk K, Zalpuri S, Prinsze FJ, Merz EM, de Kort W. Associations of health status with subsequent blood donor behavior-An alternative perspective on the Healthy Donor Effect from Donor InSight, PLoS One 2017; 12:e0186662. https://10.1371/journal.pone.0186662.

[40] Di Angelantonio E, Thompson SG, Kaptoge S, Moore C, Walker M, Armitage J, Ouwehand WH, Roberts DJ, Danesh J. Efficiency and safety of varying the frequency of whole blood donation (INTERVAL): a randomised trial of 45 000 donors, Lancet 2017; 390:2360–71. https://10.1016/s0140-6736(17)31928-1.

[41] Grau K, Vasan SK, Rostgaard K, Bialkowski W, Norda R, Hjalgrim H, Edgren G, National Heart L, Blood Institute Recipient E, Donor Evaluation S, III. No association between frequent apheresis donation and risk of fractures: a retrospective cohort analysis from Sweden, Transfusion 2017; 57:390–6. https://10.1111/trf.13907.

[42] Burgin M, Hopkins G, Moore B, Nasser J, Richardson A, Minchinton R. Serum IgG and IgM levels in new and regular long-term plasmapheresis donors, Med Lab Sci 1992; 49:265–70.

[43] Wasi S, Santowski T, Murray SA, Perrault RA, Gill P. The Canadian Red Cross plasmapheresis donor safety program: changes in plasma proteins after long-term plasmapheresis, Vox Sang 1991; 60:82–7. https://10.1111/j.1423-0410.1991.tb00879.x.

[44] Friedman BA, Schork MA, Alm SK, Jones AS, Oberman HA. Plasmapheresis-induced hemodilution and its effects on serum constituents, Transfusion 1976; 16:155–61. https://10.1046/j.1537-2995.1976.16276155110.x.

[45] Rezvan H, Ahmadi J, Mirbod V. The Iranian Blood Transfusion, Donor Safety Program: Effect of Long-term Plasmapheresis on Plasma Proteins, Iranian Journal of Medical Sciences 2003; 28:33–6.

[46] Schreiber GB, Brinser R, Rosa-Bray M, Yu ZF, Simon T. Frequent source plasma donors are not at risk of iron depletion: the Ferritin Levels in Plasma Donor (FLIPD) study, Transfusion 2018; 58:951–9. https://10.1111/trf.14489.

[47] Tran-Mi B, Storch H, Seidel K, Schulzki T, Haubelt H, Anders C, Nagel D, Siegler KE, Vogt A, Seiler D, Hellstern P. The impact of different intensities of regular donor plasmapheresis on humoral and cellular immunity, red cell and iron metabolism, and cardiovascular risk markers, Vox Sang 2004; 86:189–97. https://10.1111/j.0042-9007.2004.00408.x.

