## Appendix for "Balancing donor health and plasma collection: a systematic review of the impact of plasmapheresis frequency"

### Appendix A: Search Strategy

#### PubMed

**1. Population** (plasma donor)
"plasma donor*"[TIAB] OR “plasma donat*”[TIAB]
OR (("Plasmapheresis"[Mesh:NoExp] OR "plasmapheres*"[TIAB] OR "plasmaphaeres*"[TIAB] OR “plasma collection”[TIAB] OR “plasma withdrawal”[TIAB] OR "plasma removal"[TIAB]
OR “Blood component removal”[Mesh:NoExp] OR "apheres*"[TIAB] OR "aphaeres*"[TIAB] OR "pheres*"[TIAB] OR "phaeres*"[TIAB]
OR "Plateletpheresis"[Mesh:NoExp] OR "plateletpheres*"[TIAB] OR "plateletphaeres*"[TIAB] OR thrombocytapheres*[TIAB] OR thrombocytopheres*[TIAB] OR thrombapheres*[TIAB] OR thromboapheres*[TIAB] OR thrombophores*[TIAB] OR thrombocytaphaeres*[TIAB] OR thrombocytophaeres*[TIAB] OR thrombaphaeres*[TIAB] OR thromboaphaeres*[TIAB])
AND ("donor*"[TIAB] OR "donat*"[TIAB]))

**2. Concept** (safety)
"safety"[MESH] OR “safe*”[TIAB] OR “efficacy”[TIAB] OR “effect*”[TIAB]
OR ((“adverse”[TIAB] OR “undesirable”[TIAB] OR "side"[TIAB] OR "acute"[TIAB] OR "short-term"[TIAB] OR "long-term"[TIAB] OR "shortterm"[TIAB] OR "longterm"[TIAB] OR vital[TIAB]) AND (“reaction*”[TIAB] OR “consequence*”[TIAB] OR “event*”[TIAB] OR “outcome*”[TIAB] OR “symptom*”[TIAB]))
OR “tolerability”[TIAB] OR “harm”[TIAB] OR “complication*”[TIAB]
OR “plasmavigilance”[TIAB] OR “hemovigilance”[TIAB] OR “haemovigilance”[TIAB]
OR "donor vigilance"[TIAB]
OR "health"[TIAB] OR "monitor*"[TIAB] OR "surveillance"[TIAB]
OR "Plasmapheresis/adverse effects"[MESH] OR "Blood Component Removal/adverse effects"[Mesh]
OR "Plateletpheresis/adverse effects"[Mesh] OR "Monitoring, physiologic"[Mesh]

**3. Concept** (risk and prevention)
“Preventive Medicine”[Mesh] OR “prevention and control”[Subheading] OR “Health Promotion”[Mesh] OR “Risk Factors”[Mesh] OR “prevent*”[TIAB] OR "protect*"[TIAB] OR “risk factor”[TIAB] OR “risk factors”[TIAB] OR “Risk Assessment”[Mesh:NoExp] OR “Risk”[Mesh:NoExp] OR "donor characteristic*"[TIAB]
OR "Plasmapheresis/methods"[MESH]

#1 AND (#2 OR #3)

#### Embase

**1. Population** (plasma donor)
'plasma donor*':ti,ab OR 'plasma donat*':ti,ab
OR ((Plasmapheresis/de OR plasmapheres*:ti,ab OR plasmaphaeres*:ti,ab OR 'plasma collection':ti,ab OR 'plasma withdrawal':ti,ab OR 'plasma removal':ti,ab
OR apheresis/de OR apheres*:ti,ab OR aphaeres*:ti,ab OR pheres*:ti,ab OR phaeres*:ti,ab
OR thrombocytopheresis/de OR plateletpheres*:ti,ab OR plateletphaeres*:ti,ab OR thrombocytapheres*:ti,ab OR thrombocytopheres*:ti,ab OR thrombapheres*:ti,ab OR thromboapheres*:ti,ab OR thrombophores*:ti,ab OR thrombocytaphaeres*:ti,ab OR thrombocytophaeres*:ti,ab OR thrombaphaeres*:ti,ab OR thromboaphaeres*:ti,ab) AND (donor*:ti,ab OR donat*:ti,ab))

**2. Concept** (safety)
safety/exp OR safe*:ti,ab OR efficacy:ti,ab OR effect*:ti,ab
OR ((adverse:ti,ab OR undesirable:ti,ab OR side:ti,ab OR acute:ti,ab OR 'short-term':ti,ab OR 'long-term':ti,ab OR shortterm:ti,ab OR longterm:ti,ab OR vital:ti,ab) AND (reaction*:ti,ab OR consequence:ti,ab OR event*:ti,ab OR outcome*:ti,ab OR symptom*:ti,ab))
OR tolerability:ti,ab OR harm:ti,ab OR complication*:ti,ab
OR plasmavigilance:ti,ab OR hemovigilance:ti,ab OR haemovigilance:ti,ab
OR 'donor vigilance':ti,ab
OR health:ti,ab OR monitor*:ti,ab OR surveillance:ti,ab
OR 'physiologic monitoring'/exp

**3. Concept** (risk and prevention)

'Preventive Medicine'/exp OR 'Health Promotion'/exp OR 'Risk Factor'/exp OR prevent*:ti,ab OR protect*:ti,ab OR 'risk factor':ti,ab OR 'risk factors':ti,ab OR 'Risk Assessment'/de OR 'Risk'/de OR 'donor characteristic*':ti,ab

#1 AND (#2 OR #3)

#### Cochrane Library

**1. Population** (plasma donor)
(plasma NEXT donor*):ti,ab,kw OR (plasma NEXT donat*):ti,ab,kw
OR (([mh ^"Plasmapheresis"] OR plasmapheres*:ti,ab,kw OR plasmaphaeres*:ti,ab,kw OR "plasma collection":ti,ab,kw OR "plasma withdrawal":ti,ab,kw OR "plasma removal":ti,ab,kw
OR [mh ^"Blood component removal"] OR apheres*:ti,ab,kw OR aphaeres*:ti,ab,kw OR pheres*:ti,ab,kw OR phaeres*:ti,ab,kw
OR [mh ^"Plateletpheresis"] OR plateletpheres*:ti,ab,kw OR plateletphaeres*:ti,ab,kw OR thrombocytapheres*:ti,ab,kw OR thrombocytopheres*:ti,ab,kw OR thrombapheres*:ti,ab,kw OR thromboapheres*:ti,ab,kw OR thrombophores*:ti,ab,kw OR thrombocytaphaeres*:ti,ab,kw OR thrombocytophaeres*:ti,ab,kw OR thrombaphaeres*:ti,ab,kw OR thromboaphaeres*:ti,ab,kw)
AND (donor*:ti,ab,kw OR donat*:ti,ab,kw))

**2. Concept** (safety)
[mh "safety"] OR safe*:ti,ab,kw OR efficacy:ti,ab,kw OR effect*:ti,ab,kw
OR ((adverse:ti,ab,kw OR undesirable:ti,ab,kw OR side:ti,ab,kw OR acute:ti,ab,kw OR "short-term":ti,ab,kw OR "long-term":ti,ab,kw OR shortterm:ti,ab,kw OR longterm:ti,ab,kw OR vital:ti,ab,kw) AND (reaction*:ti,ab,kw OR consequence:ti,ab,kw
OR event*:ti,ab,kw OR outcome*:ti,ab,kw OR symptom*:ti,ab,kw))
OR tolerability:ti,ab,kw OR harm:ti,ab,kw OR complication*:ti,ab,kw
OR plasmavigilance:ti,ab,kw OR hemovigilance:ti,ab,kw OR haemovigilance:ti,ab,kw
OR "donor vigilance":ti,ab,kw
OR health:ti,ab,kw OR monitor*:ti,ab,kw OR surveillance:ti,ab,kw
OR [mh "Monitoring, Physiologic "]

**3. Concept** (risk and prevention)
[mh "Preventive Medicine"] OR [mh "Health Promotion"] OR [mh "Risk Factors"]
OR prevent*:ti,ab,kw OR protect*:ti,ab,kw OR (risk NEXT (factor OR factors)):ti,ab,kw OR [mh ^"Risk Assessment"] OR [mh ^"Risk"] OR (donor NEXT characteristic*):ti,ab,kw

#1 AND (#2 OR #3)

#### Web of Science Core Collection (Science Citation Index Expanded (SCI-EXPANDED) and Conference Proceedings Citation Index- Science (CPCI-S))

**1. Population** (plasma donor)
TS=("plasma donor*") OR TS=("plasma donat*")
OR ((TS=("plasmapheres*") OR TS=("plasmaphaeres*") OR TS=("plasma collection") OR TS=("plasma withdrawal") OR TS=("plasma removal")
OR (TS="apheres*") OR (TS="aphaeres*") OR (TS="pheres*") OR (TS="phaeres*")
OR (TS="plateletpheres*") OR (TS="plateletphaeres*") OR (TS="thrombocytapheres*") OR (TS="throm bocytopheres*") OR (TS="thrombocytepheres*") OR (TS="thrombapheres*") OR (TS="thromboapheres *") OR (TS="thrombophores*") OR (TS="thrombocytaphaeres*") OR (TS="thrombocytophaeres*")
OR (TS="thrombocytephaeres*") OR (TS="thrombaphaeres*") OR (TS="thromboaphaeres*"))
AND (TS=("donor*") OR TS=("donat*")))

**2. Concept** (safety)
TS=("safe*") OR TS=("efficacy") OR TS=("effect*")
OR ((TS=("adverse") OR TS=("undesirable") OR TS=("side") OR TS=("acute") OR TS=("short-term") OR TS=("long-term") OR TS=("shortterm") OR TS=("longterm") OR TS=("vital")) AND (TS=("reaction*") OR TS=("consequence*") OR
TS=("event*") OR TS=("outcome*") OR TS=("symptom*")))
OR TS=("tolerability") OR TS=("harm") OR TS=("complication*")
OR TS=("plasmavigilance") OR TS=("hemovigilance") OR TS=("haemovigilance")
OR TS=("donor vigilance")
OR TS=("health") OR TS=("monitor*") OR TS=("surveillance")

**3. Concept** (risk and prevention)
TS=("prevent*") OR TS=("protect*") OR TS=("risk factor") OR TS=("risk factors") OR TS=("donor characteristic*")

#1 AND (#2 OR #3)

#### CINAHL (via the Ebsco interface)

**1. Population** (plasma donor)
(TI "plasma donor*" OR AB "plasma donor*") OR (TI "plasma donat*" OR AB "plasma donat*")
OR (((MH "Plasmapheresis") OR (TI "plasmapheres*" OR AB "plasmapheres*") OR (TI "plasmaphaeres*" OR AB "plasmaphaeres*") OR (TI "plasma collection" OR AB "plasma collection") OR (TI "plasma withdrawal" OR AB "plasma withdrawal") OR (TI "plasma removal" OR AB "plasma removal")
OR (MH "Blood component removal") OR (TI "apheres*" OR AB "apheres*") OR (TI "aphaeres*" OR AB "aphaeres*") OR (TI "pheres*" OR AB "pheres*") OR (TI "phaeres*" OR AB "phaeres*")
OR (MH "Plateletpheresis") OR (TI "plateletpheres*" OR AB "plateletpheres*") OR (TI "plateletphaeres*" OR AB "plateletphaeres*") OR (TI "thrombocytapheres*" OR AB "thrombocytapheres*") OR (TI "thrombocytopheres*" OR AB "thrombocytopheres*") OR (TI "thrombapheres*" OR AB "thrombapheres*") OR (TI "thromboapheres*" OR AB "thromboapheres*") OR (TI "thrombophores*" OR AB "thrombophores*") OR (TI "thrombocytaphaeres*" OR AB "thrombocytaphaeres*") OR (TI "thrombocytophaeres*" OR AB "thrombocytophaeres*") OR (TI "thrombaphaeres*" OR AB "thrombaphaeres*") OR (TI "thromboaphaeres*" OR AB "thromboaphaeres*"))
AND ((TI "donor*" OR AB "donor*") OR (TI "donat*" OR AB "donat*")))

**2. Concept** (safety)
(MH safety+) OR (TI safe* OR AB safe*) OR (TI effect* OR AB effect*)
OR (((TI adverse OR AB adverse) OR (TI undesirable OR AB undesirable) OR (TI side OR AB side) OR (TI acute OR AB acute) OR (TI short-term OR AB short-term) OR (TI long-term OR AB long-term) OR (TI shortterm OR AB shortterm) OR (TI longterm OR AB longterm)
OR (TI vital OR AB vital)) AND ((TI reaction* OR AB reaction*) OR (TI consequence OR AB consequence) OR (TI event* OR AB event*) OR (TI outcome* OR AB outcome*) OR (TI symptom* OR AB symptom*)))
OR (TI tolerability OR AB tolerability) OR (TI harm OR AB harm) OR (TI complication* OR AB complicati on*)
OR (TI plasmavigilance OR AB plasmavigilance) OR (TI hemovigilance OR AB hemovigilance) OR (TI hae movigilance OR AB haemovigilance)
OR (TI "donor vigilance" OR AB "donor vigilance")
OR (TI health OR AB health) OR (TI monitor* OR AB monitor*) OR (TI surveillance OR AB surveillance)
OR (MH "Monitoring, physiologic"+)

**3. Concept** (risk and prevention)

(MH "Preventive Medicine"+)
OR "prevention and control[Subheading]" OR (MH "Risk Factors"+) OR (TI prevent* OR AB prevent*) OR (TI protect* OR AB protect*) OR (TI "risk factor" OR AB "risk factor") OR (TI "risk factors" OR AB "risk factors")OR (MH "Risk Assessment") OR (TI "donor characteristic*" OR AB "donor characteristic*")

#### Transfusion Evidence Library

Filter: Clinical speciality > Blood Donors
((plasma OR plasmapheresis OR plasmaphaeresis OR apheresis OR aphaeresis OR pheresis OR phaeresis OR plateletpheresis OR plateletphaeresis OR thrombocytapheresis OR thrombocytopheresis OR thrombapheresis OR thromboapheresis OR thrombophoresis OR thrombocytaphaeresis OR thrombocytophaeresis OR thrombaphaeresis OR thromboaphaeresis) AND (donor OR donors OR donation OR donations))

#### PROSPERO International Prospective Register of Systematic Reviews

((plasma OR plasmapheresis OR plasmaphaeresisOR apheresis OR aphaeresis OR pheresis OR phaeresis OR plateletpheresis OR plateletphaeresis OR thrombocytapheresis OR thrombocytopheresis OR thrombapheresis OR thromboapheresis OR thrombophoresis OR thrombocytaphaeresis OR thrombocytophaeresis OR thrombaphaeresis OR thromboaphaeresis) AND (donor OR donors OR donation OR donations))
AND (safe OR safety OR effect OR ((adverse OR undesirable OR side OR acute OR "short-term" OR "long-term" OR shortterm OR longterm) AND (reaction OR event OR outcome OR symptom OR reactions OR effects OR events OR outcomes OR symptoms)) OR tolerability OR harm OR complication OR complications OR plasmavigilance OR hemovigilance OR haemovigilance OR "donor vigilance" OR health OR monitor OR monitoring OR surveillance OR prevent OR prevention OR protect OR protection OR risk OR characteristic OR characteristics)

#### International Clinical Trials Registry Platform (trialsearch.who.int/)

((plasma OR plasmapheresis OR plasmaphaeresis OR apheresis OR aphaeresis OR pheresis OR phaeresis OR plateletpheresis OR plateletphaeresis OR thrombocytapheresis OR thrombocytopheresis OR thrombapheresis OR thromboapheresis OR thrombophoresis OR thrombocytaphaeresis OR thrombocytophaeresis OR thrombaphaeresis OR thromboaphaeresis) AND (donor OR donors OR donation OR donations))
AND (safe OR safety OR effect OR ((adverse OR undesirable OR side OR acute OR "short-term" OR "long-term" OR shortterm OR longterm) AND (reaction OR event OR outcome OR symptom OR reactions OR effects OR events OR outcomes OR symptoms)) OR tolerability OR harm OR complication OR complications OR plasmavigilance OR hemovigilance OR haemovigilance OR "donor vigilance" OR health OR monitor OR monitoring OR surveillance OR prevent OR prevention OR protect OR protection OR risk OR characteristic OR characteristics)

**Clinicaltrials.gov** (split up into 3 searches due to character limit) search in 'Other terms' field

1) (((plasma OR plasmapheresis OR apheresis OR plateletpheresis) AND (donor OR donation)))
AND
(safe OR safety OR effect OR ((adverse OR undesirable OR side OR acute OR "short-term" OR "long-term") AND (reaction OR event OR outcome OR symptom)))

2) (((plasma OR plasmapheresis OR apheresis OR plateletpheresis) AND (donor OR donation)))
AND
(plasmavigilance OR hemovigilance OR haemovigilance OR "donor vigilance" OR monitor OR monitoring OR surveillance)

3) (((plasma OR plasmapheresis OR apheresis OR plateletpheresis) AND (donor OR donation)))
AND
(tolerability OR harm OR complication OR health OR prevent OR prevention OR protect OR protection OR risk OR characteristic)

### Appendix B: Data extraction form

| **Study information** | | | | **Population** | **Intervention** | **Comparator** |
|---|---|---|---|---|---|---|
| **Author, Year, Country** | **Study sponsor** | **Financial disclosures of the co-authors** | **Study design** | | | |
| | | | | Description of the used population: number of original selected individuals, gender, age, number of individuals receiving the intervention, number of individuals in control group, nationality (when different from the country listed in the first column)<br><br>Inclusion and exclusion criteria of the studied population | Extensive description of the plasmapheresis: type of device; modalities (frequency, volume, duration) | Extensive description of the plasmapheresis: type of device; modalities (frequency, volume, duration) |

Study findings

| **Outcome** | **Comparison (Intervention vs comparator)** | **Effect size** | **#studies, #participants** | **Reference** |
|---|---|---|---|---|
| Outcome (unit) | | | | |

### Appendix C: Quality assessment

**Study limitations**

***Experimental studies***

| Author, Year | Lack of allocation concealment | Lack of blinding | Incomplete accounting of outcome events | Selective outcome reporting | Other limitations |
|---|---|---|---|---|---|
| Ciszewski, 1993 | Lack of randomization: **yes**<br>No randomization was performed<br><br>Lack of allocation concealment: **not applicable** | **unclear**, but outcomes are less likely to be influenced by a lack of blinding<br><br>Participants: **unclear**<br>No blinding reported, but outcomes are not likely to be influenced by this lack of blinding<br><br>Personnel: **unclear**<br>No blinding reported, but outcomes are not likely to be influenced by this lack of blinding<br><br>Outcome assessors: **unclear** | **yes**<br>Drop-outs are present but unclear whether the reasons were different between the groups *("a few donors were dropped from the study due to medical or technical reasons"; "Four donors had to be deferred because of protein concentrations falling below accepted limits")*<br><br>Difference in proportion of drop-outs across groups<br><br>Per protocol analysis | **yes**<br>No detailed information on differences in adverse events (donor acceptance) and albumin levels between groups | Limited compliance to plasma donor regimen (*"although they all promised to keep appointments, some were not able to do so")*: n varies between 11 and 15 for weekly group, between 12 and 30 for 14-day interval group<br><br>Sources of potential selection bias:<br>- only established donors, who probably tolerate plasmapheresis well, were included (but no quantitative information is provided on plasmapheresis history)<br>- donors with protein levels close to the lower border of accepted limits were not allowed to participate<br><br>Donor groups "relatively closely matched in gender, age and weight" at the start of the study, but no statistics to check equivalence (data not |

| | | | | | |
|---|---|---|---|---|---|
| | | | | | provided for participants included in final analysis) |
| Mortier, 2023, Belgium | Lack of randomization: **no**<br>Participants were randomly assigned to groups via a computer-generated randomization table<br><br>"The randomization procedure was suboptimal since allocation to the high-frequency/very high-frequency group was sometimes in conflict with the availability of the participant. Therefore, the donor was assigned to the first available position on | Participants: **yes (partially)**<br>"The placebo group underwent a placebo donation at the same frequency as LF (1x/month). All participants were blinded to the group selection.", although they were of course aware of the frequency at which they came to the centre<br><br>Personnel: **unclear**<br>No blinding reported, but outcomes are less | **yes**<br><br>Nine subjects (2 in the placebo group, 4 in the HF group and 3 in the VHF group) gave up before the end of the study. Withdrawal from the study was due to personal reasons or impossibility to comply with the repeated appointments.<br>Data of the drop-outs were not included in the analyses, i.e. per protocol analysis | **no** | Trial registered (NCT05815615) only after it was performed<br><br>Limited compliance to plasma donor regimen (13 subjects were not compliant with the donor regimen (by missing donations), especially in the VHF group:<br>-Group P: n = 17 (15 completed, all compliant with donor regimen)<br>-Group LF: n = 16<br>(16 completed, 15 compliant with donor regimen)<br>-Group HF: n = 20 (16 completed, 14 compliant with donor regimen)<br>-Group VHF: n = 19 (16 completed; 5 compliant with donor regimen: 6 subjects missed 1 donation, 2 missed |

| | | | | | |
|---|---|---|---|---|---|
| | the randomization list that did not conflict with his availability. The impact of this suboptimal randomization procedure was considered to be limited."<br><br>"At the start of the study, the 4 groups were not different regarding age, BMI, the amount of physical activity per week, VO2 peak and donation history."<br><br>Lack of allocation concealment: **unclear** | likely to be influenced by this lack of blinding<br><br>Outcome assessors: **unclear** | | | 2-3 donations, 2 missed 5-6 donations, 1 missed 10 donations))<br><br>Sources of potential selection bias:<br>- new and established donors were eligible, but (except for 1 new donor) only established donors, who probably tolerate plasmapheresis well, participated in the study (no quantitative information is provided on plasmapheresis history)<br><br>35 outcomes were assessed, leading to an increased rate of false positives for which no correction was applied. Sidak corrections were applied in contrast analyses to compare means.<br><br>Population limited to men aged 18-50 (average 31.4-37.7) |

**Certainty of the body of evidence for the experimental studies**

| | **Initial grading High [A]** | Downgrading due to |
|---|---|---|
| **Limitations of study design** | -1 | See table 'Study limitations' |
| **Imprecision** | -1 | Limited sample sizes |
| **Inconsistency** | 0 | |
| **Indirectness** | 0 | Limited external validity (one old study and one study with men only), population of active donors |

| | |
|---|---|
| **Publication bias** | 0 |
| **CERTAINTY (GRADE)** | **Final grading Low [C]** |

***Observational studies***

| Author, Year | Inappropriate eligibility criteria | Inappropriate methods for exposure and outcome variables | Not controlled for confounding | Incomplete or inadequate follow-up | Other limitations |
|---|---|---|---|---|---|
| Grgicevic, 1980 | **unclear**<br><br>Selection of participants limited to male student population at the University of Zagreb<br><br>Unclear what the "national criteria for blood donors" are<br><br>Unclear which total protein level was used to exclude donors.<br>In addition, seven donors were rejected due to the repeated low values of total proteins or the change | **no**<br><br>No indication of differential surveillance for outcomes in the interventions and comparator group. | **yes**<br><br>Failure of accurate measurement of all known prognostic factors (e.g. body weight, BMI, age)<br><br>Lack of adjustment for weight, baseline protein levels in statistical analysis (Student's t-test)<br><br>Subgroup analysis according to the number of plasmapheresis sessions was performed (7, 15, 35, 75, >100) for some outcomes only | **yes**<br><br>Drop-out rate was very high but similar in both groups (66% in the intervention group versus 67% in the comparator group). This loss to follow-up could potentially change the overall results (i.e. no difference) in an important way.<br><br>Follow-up period was different between intervention (2 years) and comparator group (3 years) | **yes**<br><br>In addition to the different follow-up periods, the number of donations at the final outcome assessment was also different between intervention and comparison groups (155 in the intervention group versus 138 in the comparator group)<br><br>Unclear whether participants were first-time or repeat plasma donors (i.e. no information on donation history)<br><br>For most biochemical outcomes no information is given on when they were assessed (after how many donations) |

| | | | | | |
|---|---|---|---|---|---|
| | in the electrophoresis of proteins, it was unclear to which group these donors were allocated. | | | | |
| Grgicevic, 1983 | **unclear**<br><br>Male population, unclear whether they were students<br><br>Unclear what the specific inclusion/exclusion criteria were | **no**<br><br>No indication of differential surveillance for outcomes in the interventions and comparator group. | **yes**<br><br>Lack of adjustment for weight, baseline protein levels in statistical analysis (Student's t-test)<br><br>Subgroup analysis according to the number of plasmapheresis sessions was performed (7, 15, 35, 75, >100) | **yes**<br><br>total amount of participants at baseline: unclear<br><br>Drop-out was present in both groups and there was a high variability in the number of donors at different stages of the study (problem of compliance?).<br>This loss to follow-up could potentially change the overall results (i.e. majority of outcomes no difference) in an important way. | **yes**<br><br>Subgroup analysis: cut-offs were chosen arbitrarily and specific number of plasmapheresis sessions varied between groups<br><br>Unclear whether participants were first-time or repeated plasma donors (i.e. no information on donation history) |

| | | | | | |
|---|---|---|---|---|---|
| | | | | Follow-up period in both groups was unclear | |
| Rosa-Bray 2013 | **no**<br><br>All participants were required to meet eligibility criteria for plasma donation<br><br>Potential study participants were either new plasmapheresis donors or previous donors who had not donated for at least 6 months.<br><br>Participants were between 18 and 69 years of age and met all routine plasma industry standards for weight (‡110 pounds), blood pressure, pulse, temperature, haematocrit and total protein, passed a physical | **no**<br><br>No indication of differential surveillance for outcomes in the interventions and comparator group. | **no**<br><br>Variables considered to be possible predictors for change in cholesterol levels included gender, age, weight (and therefore donation volume), race, baseline cholesterol levels, total number of donations, number of donations prior to a measurement and days between donations. These variables were added and removed in a stepwise procedure, testing the overall effect and statistical significance of each predictor. The variables baseline cholesterol level, time between donations and gender were selected as predictors in the final model as the others | **yes**<br><br>A total of 666 donors were enrolled; data from 663 donors were analysed. Two donors who completed only one donation and one donor who took cholesterol-lowering medications were excluded from analysis.<br><br>Relatively few donors (<200) completed the study. This was in part due to the inclusion of many first-time donors in the study who did not have first-hand familiarity with the donation process. Study discontinuation did not appear to be due to adverse events. Most | **yes**<br><br>Funder and lead authors (with an active role in the trial) have an (undisclosed) conflict of interest<br><br>No correction for post-hoc multiple testing<br><br>Subgroups 'days between donations': cut-offs were chosen arbitrarily.<br><br>"Participants were provided with their cholesterol results 2 months into the study, presenting a possible bias if donors began lifestyle modifications after receiving this information. However, donor activities, including diet, medication and exercise, were followed using the visit questionnaire, and no changes in these activities were observed." (no information was available in the manuscript) |

| | | | | | |
|---|---|---|---|---|---|
| | examination and provided a detailed medical history.<br><br>Both male and female donors had similar profiles, with the typical donor being in the youngest age group, of white race and weighing <200 pounds. | | showed no significant effect on cholesterol-level change.<br><br>Baseline cholesterol levels were grouped according to the recommendations of the National Cholesterol Education Program.<br><br>Caveat: Multivariable repeated measures regression model with a general estimating equations approach was used. The parameterization employed in the GEE approach may be overly simplistic.<br><br>The current model is dependent on the choice of categories, and it is possible that modelling using other categories or nonlinear continuous variables would produce different results | dropouts who provided a reason for study discontinuation gave reasons that were unrelated to plasma donation or study participation. | Relatively few donors had high baseline total (38 donors) or LDL (41 donors) cholesterol<br><br>Exact sample sizes per subgroup (per each donation frequency) were not given and were estimated based on the distribution of participants across baseline cholesterol levels, possibly resulting in deviations from the true sample sizes due to differences in distributions across baseline levels among the different frequency groups and due to rounding errors. Authors were contacted twice to obtain the correct sample sizes, but did not respond |

| | | | | | |
|---|---|---|---|---|---|
| | | | Participants also completed a questionnaire to monitor changes in diet, physical activity or the use of medications that could impact cholesterol levels. However, the authors did not provide any information on this confounding factors in the results. | | |
| Salvaggio, 1971 | **unclear**<br><br>Male prisoner population<br><br>Unclear what the specific inclusion/exclusion criteria were (if present)<br><br>Not clear whether the study was prospective or retrospective | **no**<br><br>No indication of differential surveillance for outcomes in the interventions and comparator group. | **yes**<br><br>Lack of adjustment for weight, baseline protein levels in statistical analysis<br><br>No information regarding statistical analysis | **unclear**<br><br>No drop-out reported (in all groups), but possibly a retrospective study | **yes**<br><br>No detailed/quantified information regarding the questionnaire on the frequency and severity of adverse events<br><br>Not clear when outcomes were assessed precisely |

**Certainty of the body of evidence for the observational studies**

| | **Initial grading Low** | Downgrading due to |
|---|---|---|

| | | |
|---|---|---|
| **Limitations of study design** | -1 | See table 'Study limitations' |
| **Imprecision** | -1 | Limited sample sizes |
| **Inconsistency** | 0 | |
| **Indirectness** | 0 | Limited external validity: old papers, most papers men only, most papers have populations of students (young age) or prisoners |
| **Publication bias** | 0 | |
| | | Upgrading due to |
| **Large magnitude of effect** | 0 | |
| **Dose-response gradient** | 0 | |
| **Plausible confounding** | 0 | |
| **CERTAINTY (GRADE)** | **Final grading Very low** | |

### Appendix D: Detailed synthesis of findings

#### Plasmapheresis twice per week versus three times per month versus once per month versus placebo

| Outcome | Comparison (Intervention vs comparator) | Effect size | #studies, #participants | Reference |
|---|---|---|---|---|
| Albumin [g/l] | Interaction between Group (placebo donation 1x/month (P); plasma donation 1x/month (LF); plasma donation 3x/month (HF); plasma donation 2x/week (VHF)) and Time (1 week before 1st donation (D0); 42 days after 1st donation (D42); 84 days after 1st donation (D84)) | Group*Time<br>Statistically significant:<br>p <0.0001<br><br>Group differences at D84:<br><br>HF vs LF:<br>Not statistically significant:<br>44.7±3.2 vs 43.7±2.8<br>MD: 0.97, 95%CI[-0.90;2.84][a] (p=0.31)<br><br>HF vs P:<br>Not statistically significant:<br>44.7±3.2 vs 45.9±2.7<br>MD: -1.16, 95%CI[-3.06;0.74][a] (p=0.23)<br><br>HF vs VHF:<br>Statistically significant:<br>44.7±3.2 vs 40.6±2.4<br>MD: 4.13, 95%CI[2.26;6.00][a] (p<0.0001)<br><br>LF vs P:<br>Statistically significant:<br>43.7±2.8 vs 45.9±2.7<br>MD: -2.13, 95%CI[-4.03;-0.23][a] (p=0.03)<br><br>LF vs VHF:<br>Statistically significant:<br>43.7±2.8 vs 40.6±2.4 | 1, 15 (P) vs 16 (LF) vs 16 (HF) vs 16 (VHF) § | Mortier, 2023 |

| | | | | |
|---|---|---|---|---|
| | | MD: 3.16, 95%CI[1.29;5.03][a] (p=0.001)<br><br>P vs VHF:<br>Statistically significant:<br>45.9±2.7 vs 40.6±2.4<br>MD: 5.29, 95%CI[3.39;7.19][a] (p<0.0001) | | |
| Haemoglobin [g/dl] | Interaction between Group (P vs LF vs HF vs VHF) and Time (D0, D42, D84) | Group*Time<br>Statistically significant:<br>p <0.0001<br><br>Group differences at D84:<br><br>HF vs LF:<br>Not statistically significant:<br>15.0±0.8 vs 14.5±0.8<br>MD: 0.48, 95%CI[-0.13;1.08][a] (p=0.13)<br><br>HF vs P:<br>Not statistically significant:<br>15.0±0.8 vs 15.1±0.77<br>MD: -0.14, 95%CI[-0.76;0.48][a] (p=0.66)<br><br>HF vs VHF:<br>Statistically significant:<br>15.0±0.8 vs 13.9±0.8<br>MD: 1.1, 95%CI[0.49;1.71][a] (p=0.001)<br><br>LF vs P:<br>Not statistically significant:<br>14.5±0.8 vs 15.1±0.77<br>MD: -0.61, 95%CI[-1.23;0.00][a] (p=0.054)<br><br>LF vs VHF:<br>Statistically significant:<br>14.5±0.8 vs 13.9±0.8<br>MD: 0.63, 95%CI[0.02;1.23][a] (p=0.46)<br><br>P vs VHF:<br>Statistically significant: | 1, 15 (P) vs 16 (LF) vs 16 (HF) vs 16 (VHF) § | Mortier, 2023 |

| | | | | |
|---|---|---|---|---|
| | | 15.1±0.77 vs 13.9±0.8<br>MD: 1.24, 95%CI[0.62;1.86][a] (p=0.0001) | | |
| Ferritin [µg/l] | Interaction between Group (P vs LF vs HF vs VHF) and Time (D0, D42, D84) | Group*Time<br>Statistically significant:<br>p=0.0014<br><br>Group differences at D84:<br><br>HF vs LF:<br>Statistically significant:<br>31.0±14.8 vs 74.6±66<br>MD: -43.56, 95%CI[-75.78;-11.34][a] (p=0.01)<br><br>HF vs P:<br>Statistically significant:<br>31.0±14.8 vs 82.7±55.77<br>MD: -51.67, 95%CI[-84.42;-18.91][a] (p=0.003)<br><br>HF vs VHF:<br>Not statistically significant:<br>31.0±14.8 vs 20.1±10<br>MD: 10.88, 95%CI[-21.35;43.10]¥[a] (p=0.51)<br><br>LF vs P:<br>Not statistically significant:<br>74.6±66 vs 82.7±55.77<br>MD: -8.10, 95%CI[-40.86;24.65][a] (p=0.63)<br><br>LF vs VHF:<br>Statistically significant:<br>74.6±66 vs 20.1±10<br>MD: 54.44, 95%CI[22.22;86.66][a] (p=0.001)<br><br>P vs VHF:<br>Statistically significant:<br>82.7±55.77 vs 20.1±10<br>MD: 62.54, 95%CI[29.79;95.30][a] (p=0.0003) | 1, 15 (P) vs 16 (LF) vs 16 (HF) vs 16 (VHF) | Mortier, 2023 |

| | | | | |
|---|---|---|---|---|
| C-reactive protein (CRP) [mg/l] | Interaction between Group (P vs LF vs HF vs VHF) and Time (D0, D42, D84) | Group*Time<br>Not statistically significant:<br>p=0.9484 | 1, 15 (P) vs 16 (LF) vs 16 (HF) vs 16 (VHF) § | Mortier, 2023 |
| Glycemia [mg/dl] | Interaction between Group (P vs LF vs HF vs VHF) and Time (D0, D42, D84) | Group*Time<br>Statistically significant:<br>p=0.0361<br><br>Group differences at D84:<br><br>HF vs LF:<br>Statistically significant:<br>78.6±14.8 vs 92.4±22.4<br>MD: -13.75, 95%CI[-24.69;-2.81][a] (p=0.02)<br><br>HF vs P:<br>Statistically significant:<br>78.6±14.8 vs 91.4±17.82<br>MD: -12.78, 95%CI[-23.89;-1.66][a] (p=0.03)<br><br>HF vs VHF:<br>Not statistically significant:<br>78.6±14.8 vs 79.9±14.4<br>MD: -1.25, 95%CI[-12.19;9.69][a] (p=0.82)<br><br>LF vs P:<br>Not statistically significant:<br>92.4±22.4 vs 91.4±17.82<br>MD: 0.98, 95%CI[-10.14;12.09][a] (p=0.86)<br><br>LF vs VHF:<br>Statistically significant:<br>92.4±22.4 vs 79.9±14.4<br>MD: 12.5, 95%CI[1.56;23.44][a] (p=0.03)<br><br>P vs VHF:<br>Statistically significant:<br>91.4±17.82 vs 79.9±14.4<br>MD: 11.53, 95%CI[0.41;22.64][a] (p=0.04) | 1, 15 (P) vs 16 (LF) vs 16 (HF) vs 16 (VHF) § | Mortier, 2023 |

| | | | | |
|---|---|---|---|---|
| Insulinemia [pmol/l] | Interaction between Group (P vs LF vs HF vs VHF) and Time (D0, D42, D84) | Group*Time<br>Not statistically significant:<br>p=0.4047 | 1, 15 (P) vs 16 (LF) vs 16 (HF) vs 16 (VHF) § | Mortier, 2023 |
| Glycated hemoglobin (HbA1C) [%] | Interaction between Group (P vs LF vs HF vs VHF) and Time (D0, D42, D84) | Group*Time<br>Statistically significant:<br>p=0.0007<br><br>Group differences at D84:<br><br>HF vs LF:<br>Not statistically significant:<br>5.3±0.4 vs 5.3±0.4<br>MD: -0.06, 95%CI[-0.24;0.12][a] (p=0.54)<br><br>HF vs P:<br>Not statistically significant:<br>5.3±0.4 vs 5.2±0.39<br>MD: 0.02, 95%CI[-0.16;0.21][a] (p=0.81)<br><br>HF vs VHF:<br>Not statistically significant:<br>5.3±0.4 vs 5.1±0.4<br>MD: 0.15, 95%CI[-0.03;0.34][a] (p=0.10)<br><br>LF vs P:<br>Not statistically significant:<br>5.3±0.4 vs 5.2±0.39<br>MD: 0.08, 95%CI[-0.10;0.26][a] (p=0.40)<br><br>LF vs VHF:<br>Statistically significant:<br>5.3±0.4 vs 5.1±0.4<br>MD: 0.21, 95%CI[0.03;0.39][a] (p=0.03)<br><br>P vs VHF:<br>Not statistically significant:<br>5.2±0.39 vs 5.1±0.4<br>MD: 0.13, 95%CI[-0.05;0.32][a] (p=0.17) | 1, 15 (P) vs 16 (LF) vs 16 (HF) vs 16 (VHF) § | Mortier, 2023 |

| | | | | |
|---|---|---|---|---|
| Creatine kinase (CK) [U/l] | Interaction between Group (P vs LF vs HF vs VHF) and Time (D0, D42, D84) | Group*Time<br>Not statistically significant:<br>p=0.4489 | 1, 15 (P) vs 16 (LF) vs 16 (HF) vs 16 (VHF) § | Mortier, 2023 |
| Total cholesterol [mg/dl] | Interaction between Group (P vs LF vs HF vs VHF) and Time (D0, D42, D84) | Group*Time<br>Not statistically significant:<br>p=0.3901 | 1, 15 (P) vs 16 (LF) vs 16 (HF) vs 16 (VHF) § | Mortier, 2023 |
| IgA [g/l] | Interaction between Group (P vs LF vs HF vs VHF) and Time (D0, D42, D84) | Group*Time<br>Statistically significant:<br>p<0.0001<br><br>Group differences at D84:<br><br>HF vs LF:<br>Not statistically significant:<br>2.10±0.64 vs 1.89±0.88<br>MD: 0.21, 95%CI[-0.40;0.82][a] (p=0.51)<br><br>HF vs P:<br>Not statistically significant:<br>2.10±0.64 vs 2.34±1.05<br>MD: -0.24, 95%CI[-0.86;0.38][a] (p=0.45)<br><br>HF vs VHF:<br>Not statistically significant:<br>2.10±0.64 vs 2.06±0.88<br>MD: 0.04, 95%CI[-0.57;0.65][a] (p=0.90)<br><br>LF vs P:<br>Not statistically significant:<br>1.89±0.88 vs 2.34±1.05<br>MD: -0.45, 95%CI[-1.07;0.17][a] (p=0.16)<br><br>LF vs VHF:<br>Not statistically significant:<br>1.89±0.88 vs 2.06±0.88<br>MD: -0.17, 95%CI[-0.78;0.44][a] (p=0.59)<br><br>P vs VHF: | 1, 15 (P) vs 16 (LF) vs 16 (HF) vs 16 (VHF) § | Mortier, 2023 |

| | | | | |
|---|---|---|---|---|
| | | Not statistically significant:<br>2.34±1.05 vs 2.06±0.88<br>MD: 0.28, 95%CI[-0.34;0.90][a] (p=0.38) | | |
| IgG [g/l] | Interaction between Group (P vs LF vs HF vs VHF) and Time (D0, D42, D84) | Group*Time<br>Statistically significant:<br>p<0.0001<br><br>Group differences at D84:<br><br>HF vs LF:<br>Not statistically significant:<br>8.43±1.52 vs 8.7±2.2<br>MD: -0.27, 95%CI[-1.54;1.00][a] (p=0.68)<br><br>HF vs P:<br>Statistically significant:<br>8.43±1.52 vs 10.59±1.86<br>MD: -2.16, 95%CI[-3.45;-0.87][a] (p=0.001)<br><br>HF vs VHF:<br>Statistically significant:<br>8.43±1.52 vs 5.73±1.4<br>MD: 2.7, 95%CI[1.43;3.97][a] (p<0.0001)<br><br>LF vs P:<br>Statistically significant:<br>8.7±2.2 vs 10.59±1.86<br>MD: -1.89, 95%CI[-3.18;-0.61][a] (p=0.005)<br><br>LF vs VHF:<br>Statistically significant:<br>8.7±2.2 vs 5.73±1.4<br>MD: 2.97, 95%CI[1.70;4.24][a] (p<0.0001)<br><br>P vs VHF:<br>Statistically significant:<br>10.59±1.86 vs 5.73±1.4<br>MD: 4.86, 95%CI[3.57;6.15][a] (p<0.0001) | 1, 15 (P) vs 16 (LF) vs 16 (HF) vs 16 (VHF) § | Mortier, 2023 |

| | | | | |
|---|---|---|---|---|
| IgM [g/l] | Interaction between Group (P vs LF vs HF vs VHF) and Time (D0, D42, D84) | Group*Time<br>Statistically significant:<br>p<0.0001<br><br>Group differences at D84:<br><br>HF vs LF:<br>Not statistically significant:<br>0.73±0.32 vs 0.82±0.32<br>MD: -0.10, 95%CI[-0.33;0.14][a] (p=0.42)<br><br>HF vs P:<br>Not statistically significant:<br>0.73±0.32 vs 0.89±0.43<br>MD: -0.17, 95%CI[-0.41;0.07][a] (p=0.18)<br><br>HF vs VHF:<br>Not statistically significant:<br>0.73±0.32 vs 0.66±0.24<br>MD: 0.07, 95%CI[-0.17;0.30][a] (p=0.58)<br><br>LF vs P:<br>Not statistically significant:<br>0.82±0.32 vs 0.89±0.43<br>MD: -0.07, 95%CI[-0.31;0.17][a] (p=0.57)<br><br>LF vs VHF:<br>Not statistically significant:<br>0.82±0.32 vs 0.66±0.24<br>MD: 0.16, 95%CI[-0.07;0.40][a] (p=0.18)<br><br>P vs VHF:<br>Not statistically significant:<br>0.89±0.43 vs 0.66±0.24<br>MD: 0.23, 95%CI[-0.01;0.47][a] (p=0.06) | 1, 15 (P) vs 16 (LF) vs 16 (HF) vs 16 (VHF) § | Mortier, 2023 |
| Systolic blood pressure [mmHg] | Interaction between Group (P vs LF vs HF vs VHF) and Time (D0, D42, D84) | Group*Time<br>Not statistically significant:<br>p = 0.1499 | 1, 15 (P) vs 16 (LF) vs 16 (HF) vs 16 (VHF) § | Mortier, 2023 |

| | | | | |
|---|---|---|---|---|
| Diastolic blood pressure [mmHg] | Interaction between Group (P vs LF vs HF vs VHF) and Time (D0, D42, D84) | Group*Time<br>Not statistically significant:<br>p=0.9039 | 1, 15 (P) vs 16 (LF) vs 16 (HF) vs 16 (VHF) § | Mortier, 2023 |
| Body mass [kg] | Interaction between Group (P vs LF vs HF vs VHF) and Time (D0, D42, D84) | Group*Time<br>Not statistically significant:<br>p=0.3678 | 1, 15 (P) vs 16 (LF) vs 16 (HF) vs 16 (VHF) § | Mortier, 2023 |
| BMI [kg/m$^2$] | Interaction between Group (P vs LF vs HF vs VHF) and Time (D0, D42, D84) | Group*Time<br>Not statistically significant:<br>p=0.3311 | 1, 15 (P) vs 16 (LF) vs 16 (HF) vs 16 (VHF) § | Mortier, 2023 |
| Bone mineral content (BMC) [kg] | Interaction between Group (P vs LF vs HF vs VHF) and Time (D0, D42, D84) | Group*Time<br>Not statistically significant:<br>p=0.0803 | 1, 15 (P) vs 16 (LF) vs 16 (HF) vs 16 (VHF) § | Mortier, 2023 |
| Fat mass [kg] | Interaction between Group (P vs LF vs HF vs VHF) and Time (D0, D42, D84) | Group*Time<br>Not statistically significant:<br>p=0.7008 | 1, 15 (P) vs 16 (LF) vs 16 (HF) vs 16 (VHF) § | Mortier, 2023 |
| Fat-free mass (FFM) [kg] | Interaction between Group (P vs LF vs HF vs VHF) and Time (D0, D42, D84) | Group*Time<br>Not statistically significant:<br>p=0.7596 | 1, 15 (P) vs 16 (LF) vs 16 (HF) vs 16 (VHF) § | Mortier, 2023 |
| FFM + BMC [kg] | Interaction between Group (P vs LF vs HF vs VHF) and Time (D0, D42, D84) | Group*Time<br>Not statistically significant:<br>p=0.7919 | 1, 15 (P) vs 16 (LF) vs 16 (HF) vs 16 (VHF) § | Mortier, 2023 |
| % fat | Interaction between Group (P vs LF vs HF vs VHF) and Time (D0, D42, D84) | Group*Time<br>Not statistically significant:<br>p=0.75 | 1, 15 (P) vs 16 (LF) vs 16 (HF) vs 16 (VHF) § | Mortier, 2023 |
| Lactate 190W [mmol/l] | Interaction between Group (P vs LF vs HF vs VHF) and Time (D0, D42, D84) | Group*Time<br>Not statistically significant:<br>p=0.9188 | 1, 15 (P) vs 16 (LF) vs 16 (HF) vs 16 (VHF) § | Mortier, 2023 |
| Lactate post [mmol/l] | Interaction between Group (P vs LF vs HF vs VHF) and Time (D0, D42, D84) | Group*Time<br>Not statistically significant:<br>p=0.8807 | 1, 15 (P) vs 16 (LF) vs 16 (HF) vs 16 (VHF) § | Mortier, 2023 |
| Heart rate max (HRmax) [bpm] | Interaction between Group (P vs LF vs HF vs VHF) and Time (D0, D42, D84) | Group*Time<br>Not statistically significant:<br>p=0.9383 | 1, 15 (P) vs 16 (LF) vs 16 (HF) vs 16 (VHF) § | Mortier, 2023 |
| Ventilation max (VEmax) [l/min] | Interaction between Group (P vs LF vs HF vs VHF) and Time (D0, D42, D84) | Group*Time<br>Not statistically significant:<br>p=0.8645 | 1, 15 (P) vs 16 (LF) vs 16 (HF) vs 16 (VHF) § | Mortier, 2023 |

| | | | | |
|---|---|---|---|---|
| Peak oxygen consumption ($VO_2$ peak) [ml/(min*kg)] | Interaction between Group (P vs LF vs HF vs VHF) and Time (D0, D42, D84) | Group*Time<br>Not statistically significant:<br>p=0.7932 | 1, 15 (P) vs 16 (LF) vs 16 (HF) vs 16 (VHF) § | Mortier, 2023 |
| Oxygen pulse [ml/(beat*kg)] | Interaction between Group (P vs LF vs HF vs VHF) and Time (D0, D42, D84) | Group*Time<br>Not statistically significant:<br>p=0.5445 | 1, 15 (P) vs 16 (LF) vs 16 (HF) vs 16 (VHF) § | Mortier, 2023 |
| Haematoma [rate per 50 donations; total number of occurrences] | Group (P vs LF vs HF vs VHF) | P: 0/50; 0 occurrences<br>LF: 0/50; 0 occurrences<br>HF: 1.08/50; 3 occurrences (in 3 donors)<br>VHF: 0.28/50; 2 occurrences (in 1 donor) | 1, 15 (P) vs 16 (LF) vs 16 (HF) vs 16 (VHF) § | Mortier, 2023 |
| Vasovagal reactions (without loss of consciousness) [rate per 50 donations; total number of occurrences] | Group (P vs LF vs HF vs VHF) | P: 0/50; 0<br>LF: 0/50; 0<br>HF: 0.36/50; 1 occurrence<br>VHF: 0.57/50; 4 occurrences (in 3 donors) | 1, 15 (P) vs 16 (LF) vs 16 (HF) vs 16 (VHF) § | Mortier, 2023 |
| Anaemia [rate per 50 donations; total number of occurrences] | Group (P vs LF vs HF vs VHF) | P: 0/50; 0 occurrences<br>LF: 0/50; 0 occurrences<br>HF: 0/50; 0 occurrences<br>VHF: 0.71/50; 5 occurrences (in 4 donors) | 1, 15 (P) vs 16 (LF) vs 16 (HF) vs 16 (VHF) § | Mortier, 2023 |
| Other adverse events[b] [rate per 50 donations; total number of occurrences] | Group (P vs LF vs HF vs VHF) | P: 0/50; 0 occurrences<br>LF: 0/50; 0 occurrences<br>HF: 0/50; 0 occurrences<br>VHF: 0/50; 0 occurrences | 1, 15 (P) vs 16 (LF) vs 16 (HF) vs 16 (VHF) § | Mortier, 2023 |

Mean ± SD, MD: mean difference, SD: standard deviation

* Calculations (MD, 95%CI) done by the reviewer(s) using Review Manager software

§ Imprecision (limited sample size)

[a]Means and standard deviations of the means were extracted from Tables 1-3 of the paper by Mortier (2023), only if the Group*Time interaction effect was significant (as evidenced by the p-values in Table S1 of the paper). MDs and standard errors of the MDs were taken from supplemental information (statistical reports) provided by the study authors via personal communication and were subsequently used to calculate the 95% CIs using Excel.

[b]Adverse events were registered by Mortier (2023) in accordance with the ISBT classification tool, with the notable exception that citrate reactions were not considered since donors received preventive calcium supplements.

#### Weekly versus biweekly plasmapheresis

| Outcome | Comparison (Intervention vs comparator) | Effect size | #studies, #participants | Reference |
|---|---|---|---|---|
| Total serum protein concentration [g/l] (mean±std) | Weekly vs 14-day intervals plasma donation | *At 1st month*<br>Not statistically significant:<br>69.4±2.9 vs 69.6±3.5<br>MD: -0.20, 95%CI [-2.13;1.73]<br>(p=0.84)* | 1, 15 vs 30 § | Ciszewski, 1993 |
| | | *At 2nd month*<br>Not statistically significant:<br>67.0±3.6 vs 67.5±3.6<br>MD: -0.50, 95%CI [-2.96;1.96]<br>(p=0.69)* | 1, 12 vs 26 § | |
| | | *At 3rd month*<br>Statistically significant:<br>64.6±2.6 vs 67.9±3.7<br>MD: -3.30, 95%CI [-5.57;-1.03]<br>(p=0.004)*<br>*in favour of biweekly plasma donation* | 1, 14 vs 16 § | |
| | | *At 4th month*<br>Not statistically significant:<br>65.0±3.7 vs 67.7±4.6<br>MD: -2.70, 95%CI [-5.99;0.59]<br>(p=0.11)* | 1, 13 vs 12 § | |
| | | *At 5th month*<br>Not statistically significant:<br>65.5±3.0 vs 67.2±4.9<br>MD: -1.70, 95%CI [-4.75;1.35]<br>(p=0.27)* | 1, 11 vs 15 § | |
| | | *At 6th month*<br>Statistically significant:<br>66.2±3.0 vs 69.5±3.0<br>MD: -3.30, 95%CI [-5.35;-1.25]<br>(p=0.002)*<br>*in favour of biweekly plasma donation* | 1, 14 vs 20 § | |

| | | | | |
|---|---|---|---|---|
| Immunoglobulin G levels [g/l] (mean±std) | Weekly vs 14-day intervals plasma donation | *At 1st month*<br>Not statistically significant:<br>8.7±1.6 vs 9.5±2.3<br>MD: -0.80, 95%CI [-1.95;0.35]<br>(p=0.17)* | 1, 15 vs 30 § | Ciszewski, 1993 |
| | | *At 2nd month*<br>Not statistically significant:<br>9.3±2.0 vs 10.4±2.0<br>MD: -1.10, 95%CI [-2.47;0.27]<br>(p=0.12)* | 1, 12 vs 26 § | |
| | | *At 3rd month*<br>Not statistically significant:<br>8.7±1.8 vs 9.9±2.6<br>MD: -1.20, 95%CI [-2.78;0.38]<br>(p=0.14)* | 1, 14 vs 16 § | |
| | | *At 4th month*<br>Not statistically significant:<br>8.7±1.5 vs 9.7±1.8<br>MD: -1.00, 95%CI [-2.30;0.30]<br>(p=0.13)* | 1, 13 vs 12 § | |
| | | *At 5th month*<br>Not statistically significant:<br>8.7±1.8 vs 9.7±1.9<br>MD: -1.00, 95%CI [-2.43;0.43]<br>(p=0.17)* | 1, 11 vs 15 § | |
| | | *At 6th month*<br>Not statistically significant:<br>8.8±2.0 vs 9.8±1.9<br>MD: -1.00, 95%CI [-2.34;-0.34]<br>(p=0.14)* | 1, 14 vs 20 § | |
| Immunoglobulin A levels [g/l] (mean±std) | Weekly vs 14-day intervals plasma donation | *At 1st month*<br>Not statistically significant:<br>1.65±0.64 vs 1.99±0.84<br>MD: -0.34, 95%CI [-0.78;0.10]<br>(p=0.13)* | 1, 15 vs 30 § | Ciszewski, 1993 |
| | | *At 2nd month*<br>Not statistically significant:<br>1.92±0.82 vs 2.21±0.86<br>MD: -0.29, 95%CI [-0.86;0.28] | 1, 12 vs 26 § | |

| | | | | |
|---|---|---|---|---|
| | | (p=0.32)* | | |
| | | *At 3rd month*<br>Not statistically significant:<br>1.84±0.8 vs 2.23±0.96<br>MD: -0.39, 95%CI [-1.02;0.24]<br>(p=0.22)* | 1, 14 vs 16 § | |
| | | *At 4th month*<br>Not statistically significant:<br>1.86±0.92 vs 2.1±0.98<br>MD: -0.24, 95%CI [-0.99;0.51]<br>(p=0.53)* | 1, 13 vs 12 § | |
| | | *At 5th month*<br>Not statistically significant:<br>2.1±0.86 vs 2.24±1.0<br>MD: -0.14, 95%CI [-0.86;0.58]<br>(p=0.70)* | 1, 11 vs 15 § | |
| | | *At 6th month*<br>Not statistically significant:<br>1.94±0.84 vs 2.26±0.97<br>MD: -0.32, 95%CI [-0.93;0.29]<br>(p=0.31)* | 1, 14 vs 20 § | |
| Immunoglobulin M levels [g/l] (mean±std) | Weekly vs 14-day intervals plasma donation | *At 1st month*<br>Not statistically significant:<br>1.69±0.82 vs 2.10±1.00<br>MD: -0.41, 95%CI [-0.96;0.14]<br>(p=0.14)* | 1, 15 vs 30 § | Ciszewski, 1993 |
| | | *At 2nd month*<br>Not statistically significant:<br>1.93±0.91 vs 2.20±1.00<br>MD: -0.27, 95%CI [-0.91;0.37]<br>(p=0.41)* | 1, 12 vs 26 § | |
| | | *At 3rd month*<br>Not statistically significant:<br>1.69±0.79 vs 2.30±0.95<br>MD: -0.61, 95%CI [-1.23;0.01]<br>(p=0.05)* | 1, 14 vs 16 § | |
| | | *At 4th month*<br>Not statistically significant:<br>1.65±0.70 vs 2.00±1.10 | 1, 13 vs 12 § | |

| | | | |
|---|---|---|---|
| | | MD: -0.35, 95%CI [-1.08;0.38] (p=0.35)* | |
| | | *At 5th month*<br>Not statistically significant:<br>1.77±0.93 vs 1.99±0.80<br>MD: -0.22, 95%CI [-0.90;0.46] (p=0.53)* | 1, 11 vs 15 § |
| | | *At 6th month*<br>Not statistically significant:<br>1.73±0.84 vs 1.98±0.87<br>MD: -0.25, 95%CI [-0.83;0.33] (p=0.40)* | 1, 14 vs 20 § |

Mean ± SD, MD: mean difference, SD: standard deviation
* Calculations (MD, 95%CI) done by the reviewer(s) using Review Manager software
§ Imprecision (limited sample size)

#### Three times per two weeks versus (less than) once per week

| Outcome | Comparison (Intervention vs comparator) | Effect size | #studies, #participants | Reference |
|---|---|---|---|---|
| Total serum protein concentration [g/l] (mean±std) | 3x/2 weeks vs 1x/week | Not statistically significant:<br>66.4±3.4 vs 66.6±3.8<br>MD: -0.20, 95%CI<br>[-1.93;1.53] (p=0.82)* | 1, 34 vs 33 § | Grgicevic, 1980 |
| Immunoglobulin G levels [g/l] (mean±std) | 3x/2 weeks vs 1x/week | Not statistically significant:<br>12.77±4 vs 13.41±2.83<br>MD: -0.64, 95%CI<br>[-2.30;1.02] (p=0.45)* | 1, 34 vs 33 § | Grgicevic, 1980 |
| Immunoglobulin A levels [g/l] (mean±std) | 3x/2 weeks vs 1x/week | Not statistically significant:<br>1.71±0.75 vs 1.69±0.5<br>MD: 0.02, 95%CI<br>[-0.28;0.32] (p=0.90)* | 1, 34 vs 33 § | Grgicevic, 1980 |
| Immunoglobulin M levels [g/l] (mean±std) | 3x/2 weeks vs 1x/week | Not statistically significant:<br>1.12±0.47 vs 1.18±0.6<br>MD: -0.06, 95%CI<br>[-0.32;0.20] (p=0.65)* | 1, 34 vs 33 § | Grgicevic, 1980 |
| Albumins [g/l] | 3x/2 weeks vs 1x/week | Not statistically significant:<br>39.8±2.8 vs 39.9±2.8<br>MD: -0.10, 95%CI<br>[-1.44;1.24] (p=0.88)* | 1, 34 vs 33 § | Grgicevic, 1980 |
| Electrophoresis albumin [%] | 3x/2 weeks vs 1x/week | Not statistically significant:<br>60.04±2.37 vs 60.09±2.8<br>MD: -0.05, 95%CI<br>[-1.29;1.19] (p=0.94)* | 1, 34 vs 33 § | Grgicevic, 1980 |
| Globulins [g/l] | 3x/2 weeks vs 1x/week | Not statistically significant:<br>26.6±1.9 vs 26.7±2.6<br>MD: -0.10, 95%CI<br>[-1.19;0.99] (p=0.86)* | 1, 34 vs 33 § | Grgicevic, 1980 |
| Alpha 1 globulins [g/l] | 3x/2 weeks vs 1x/week | *After 7 donations*<br>Not statistically significant:<br>2.20±0.28 vs 2.18±0.28<br>MD: 0.02, 95%CI [-0.11;0.15]<br>(p=0.77)* | 1, 34 vs 33 § | Grgicevic, 1980 |

| | | | | |
|---|---|---|---|---|
| | | *After 15 donations*<br>Not statistically significant:<br>2.31±0.36 vs 2.23±0.40<br>MD: 0.08, 95%CI [-0.10;0.26]<br>(p=0.39)* | | |
| | | *After 35 donations*<br>Not statistically significant:<br>2.29±0.33 vs 2.24±0.33<br>MD: 0.05, 95%CI [-0.11;0.21]<br>(p=0.54)* | | |
| | | *After 75 donations*<br>Not statistically significant:<br>2.34±0.33 vs 2.32±0.28<br>MD: 0.02, 95%CI [-0.13;0.17]<br>(p=0.79)* | | |
| | | *After 155 versus 138 donations*<br>Not statistically significant:<br>2.25±0.3 vs 2.3±0.28<br>MD: -0.05, 95%CI [-0.19;0.09]<br>(p=0.48)* | | |
| Alpha 1 globulins [%] | 3x/2 weeks vs 1x/week | Not statistically significant:<br>3.37±0.39 vs 3.43±0.4<br>MD: -0.06, 95%CI<br>[-0.25;0.13] (p=0.53)* | 1, 34 vs 33 § | Grgicevic, 1980 |
| Alpha 2 globulins [g/l] | 3x/2 weeks vs 1x/week | *After 7 donations*<br>Not statistically significant:<br>5.39±0.67 vs 5.33±0.60<br>MD: 0.06, 95%CI [-0.24;0.36]<br>(p=0.70)* | 1, 34 vs 33 § | Grgicevic, 1980 |
| | | *After 15 donations*<br>Not statistically significant:<br>5.50±0.50 vs 5.57±0.60<br>MD: -0.07, 95%CI [-0.33;0.19]<br>(p=0.60)* | | |
| | | *After 35 donations*<br>Not statistically significant:<br>5.68±0.80 vs 5.43±0.78<br>MD: 0.25, 95%CI [-0.13;0.63]<br>(p=0.20)* | | |

| | | | | |
|---|---|---|---|---|
| | | *After 75 donations*<br>Not statistically significant:<br>5.79±0.70 vs 5.78±0.80<br>MD: 0.01, 95%CI [-0.35;0.37]<br>(p=0.96)* | | |
| | | *After 155 versus 138 donations*<br>Not statistically significant:<br>5.70±0.64 vs 5.72±0.60<br>MD: -0.02, 95%CI [-0.32;0.28]<br>(p=0.89)* | | |
| Alpha 2 globulins [%] | 3x/2 weeks vs 1x/week | Not statistically significant:<br>8.3±1.19 vs 8.56±0.93<br>MD: -0.26, 95%CI<br>[-0.77;0.25] (p=0.32)* | 1, 34 vs 33 § | Grgicevic, 1980 |
| Beta globulins [g/l] | 3x/2 weeks vs 1x/week | *After 7 donations*<br>Not statistically significant:<br>6.38±0.51 vs 6.30±0.50<br>MD: 0.08, 95%CI [-0.16;0.32]<br>(p=0.52)* | 1, 34 vs 33 § | Grgicevic, 1980 |
| | | *After 15 donations*<br>Not statistically significant:<br>6.29±0.59 vs 6.33±0.43<br>MD: -0.04, 95%CI [-0.29;0.21]<br>(p=0.75)* | | |
| | | *After 35 donations*<br>Not statistically significant:<br>6.40±0.54 vs 6.41±0.45<br>MD: -0.01, 95%CI [-0.25;0.23]<br>(p=0.93)* | | |
| | | *After 75 donations*<br>Not statistically significant:<br>6.66±0.69 vs 6.49±0.83<br>MD: 0.17, 95%CI [-0.20;0.54]<br>(p=0.36)* | | |
| | | *After >100 donations (155 versus 138)*<br>Not statistically significant:<br>6.81±0.60 vs 6.63±0.80<br>MD: 0.18, 95%CI [-0.16;0.52]<br>(p=0.30)* | | |

| | | | | |
|---|---|---|---|---|
| Beta globulins [g/l] | 3x/2 weeks vs 1x/week | Not statistically significant:<br>10.12±0.86 vs 9.86±1.03<br>MD: 0.26, 95%CI<br>[-0.2;0.72] (p=0.26)* | 1, 34 vs 33 § | Grgicevic, 1980 |
| Gamma globulins [%] | 3x/2 weeks vs 1x/week | Not statistically significant:<br>18.17±1.81 vs 17.81±2.28<br>MD: 0.36, 95%CI<br>[-0.63;1.35] (p=0.47)* | 1, 34 vs 33 § | Grgicevic, 1980 |
| Alpha-1-antitrypsin [g/l] | 3x/2 weeks vs 1x/week | Not statistically significant:<br>2.01±0.38 vs 2.12±0.51<br>MD: -0.11, 95%CI<br>[-0.33;0.11] (p=0.32)* | 1, 34 vs 32 § | Grgicevic, 1980 |
| Alpha-1-antitrypsin [%] | 3x/2 weeks vs < 1x/week | *After 1 donation (1 vs 1)*<br>Not statistically significant:<br>94.5±10.2 vs 94.6±10.3 λ<br>MD: -0.10, 95%CI [-4.71;4.51]<br>(p=0.97)* | 1, 38 vs 38 § | Grgicevic, 1983 |
| | | *After 7 donations (7 vs 7)*<br>Not statistically significant:<br>101.6±21.1 vs 108.0±22.0 λ<br>MD: -6.40, 95%CI [-22.67;9.87]<br>(p=0.44)* | 1, 11 vs 17 § | |
| | | *After 15 donations (15 vs 15)*<br>Not statistically significant:<br>95.9±13.5 vs 105.5±28.0 λ<br>MD: -9.60, 95%CI [-23.07;3.87]<br>(p=0.16)* | 1, 10 vs 27 § | |
| | | *After 35 donations (35 vs 35)*<br>Not statistically significant:<br>104.3±19.7 vs 100.5±22.0 λ<br>MD: 3.80, 95%CI [-5.34;12.94]<br>(p=0.42)* | 1, 39 vs 41 § | |
| | | *After 75 donations (75 vs 75)*<br>Not statistically significant:<br>103.6±18.1 vs 103.4±22.3 λ<br>MD: 0.20, 95%CI [-9.02;9.42]<br>(p=0.97)* | 1, 31 vs 43 § | |
| | | *After >100 donations (156 vs 138)*<br>Not statistically significant: | 1, 36 vs 34 § | |

| | | | | |
|---|---|---|---|---|
| | | 93.4±12.2 vs 95.9±17.7 λ<br>MD: -2.50, 95%CI [-9.66;4.66]<br>(p=0.49)* | | |
| Alpha-2-macroglobulin [g/l] | 3x/2 weeks vs 1x/week | Not statistically significant:<br>2.27±0.59 vs 2.37±0.55<br>MD: -0.1, 95%CI<br>[-0.38;0.18] (p=0.48)* | 1, 34 vs 32 § | Grgicevic, 1980 |
| Alpha-2-macroglobulin [%] | 3x/2 weeks vs < 1x/week | *After 1 donation (1 vs 1)*<br>Not statistically significant:<br>100.0±13.7 vs 100.3±13.8 λ<br>MD: -0.30, 95%CI [-6.48;5.88]<br>(p=0.92)* | 1, 38 vs 38 § | Grgicevic, 1983 |
| | | *After 7 donations (7 vs 7)*<br>Not statistically significant:<br>110.3±21.7 vs 105.7±19.9 λ<br>MD: 4.6, 95%CI [-10.52;19.72]<br>(p=0.55)* | 1, 13 vs 17 § | |
| | | *After 15 donations (15 vs 15)*<br>Not statistically significant:<br>104.8±15.8 vs 105.2±22.1 λ<br>MD: -0.40, 95%CI [-12.62;11.82]<br>(p=0.95)* | 1, 12 vs 27 § | |
| | | *After 35 donations (35 vs 35)*<br>Not statistically significant:<br>100.7±19.4 vs 100.4±17.8 λ<br>MD: 0.30, 95%CI [-7.83;8.43]<br>(p=0.94)* | 1, 39 vs 42 § | |
| | | *After 75 donations (75 vs 75)*<br>Not statistically significant:<br>102.6±15.7 vs 102.5±18.3 λ<br>MD: 0.10, 95%CI [-7.56;7.76]<br>(p=0.98)* | 1, 33 vs 43 § | |
| | | *After >100 donations (156 vs 138)*<br>Not statistically significant:<br>105.4±17.2 vs 98.6±16.6 λ<br>MD: 6.80, 95%CI [-1.12;14.72]<br>(p=0.09)* | 1, 36 vs 34 § | |
| Anti-thrombin III [g/l] | 3x/2 weeks vs 1x/week | Not statistically significant:<br>0.3±0.06 vs 0.31±0.05 | 1, 34 vs 32 § | Grgicevic, 1980 |

| | | | | |
|---|---|---|---|---|
| | | MD: -0.01, 95%CI<br>[-0.04;0.02] (p=0.46)* | | |
| Anti-thrombin III [%] | 3x/2 weeks vs < 1x/week | *After 1 donation (1 vs 1)*<br>Not statistically significant:<br>103.9±16.0 vs 104.0±15.7 λ<br>MD: -0.10, 95%CI [-7.23;7.03]<br>(p=0.98)* | 1, 38 vs 38 § | Grgicevic, 1983 |
| | | *After 7 donations (7 vs 7)*<br>Not statistically significant:<br>106.6±16.1 vs 96.0±15.3 λ<br>MD: 10.60, 95%CI [-0.78;21.98]<br>(p=0.07)* | 1, 13 vs 17 § | |
| | | *After 15 donations (14 vs 16)*<br>Not statistically significant:<br>106.4±16.5 vs 103.8±26.1 λ<br>MD: 2.6, 95%CI [-10.97;16.17]<br>(p=0.71)* | 1, 12 vs 27 § | |
| | | *After 35 donations (34 vs 34)*<br>Not statistically significant:<br>107.3±20.6 vs 102±24.1 λ<br>MD: 5.30, 95%CI [-4.44;15.04]<br>(p=0.29)* | 1, 39 vs 42 § | |
| | | *After 75 donations (78 vs 76)*<br>Not statistically significant:<br>100.3±18.4 vs 100.4±16.5 λ<br>MD: -0.10, 95%CI [-8.12;7.92]<br>(p=0.98)* | 1, 33 vs 42 § | |
| | | *After >100 donations (156 vs 138)*<br>Not statistically significant:<br>102.5±17.2 vs 100.4±15.8 λ<br>MD: 2.10, 95%CI [-5.63;9.83]<br>(p=0.59)* | 1, 36 vs 34 § | |
| Plasminogen [g/l] | 3x/2 weeks vs 1x/week | Not statistically significant:<br>0.12±0.03 vs 0.12±0.04<br>MD: 0.0, 95%CI<br>[-0.02;0.02] (p=1.00)* | 1, 34 vs 32 § | Grgicevic, 1980 |
| Plasminogen [%] | 3x/2 weeks vs < 1x/week | After 1 session (1 vs 1):<br>Not statistically significant:<br>99.02±24.96 vs 98.77±24.77 λ | 1, 38 vs 38 § | Grgicevic, 1983 |

| | | | | |
|---|---|---|---|---|
| | | MD: 0.25, 95%CI<br>[-10.94;11.43] (p=0.97)* | | |
| | | After 7 sessions (7 vs 7):<br>Statistically significant:<br>79.10±22.62 vs 96.56±12.60 λ<br>MD: -17.46, 95%CI<br>[-31.14;-3.78] (p=0.01)*<br>*In favour of plasma donation <1x/week* | 1, 13 vs 17 § | |
| | | After 15 sessions (14 vs 16):<br>Not statistically significant:<br>92.25±22.75 vs 95.94±21.39 λ<br>MD: -3.69, 95%CI<br>[-18.88;11.50] (p=0.63)* | 1, 12 vs 27 § | |
| | | After 35 sessions (34 vs 34):<br>Not statistically significant:<br>95.08±24.10 vs 96.93±23.30 λ<br>MD: -1.84, 95%CI<br>[-12.24;8.55] (p=0.73)* | 1, 39 vs 41 § | |
| | | After 75 sessions (78 vs 76):<br>Not statistically significant:<br>92.13±20.47 vs 98.03±21.27 λ<br>MD: -5.90, 95%CI<br>[-15.35;3.54] (p=0.22)* | 1, 33 vs 43 § | |
| | | After >100 sessions (156 vs 138):<br>Not statistically significant:<br>92.99±22.93 vs 97.30±26.25 λ<br>MD: -4.30, 95%CI<br>[-16.05;7.44] (p=0.47)* | 1, 35 vs 33 § | |
| Fibrinogen [g/l] | 3x/2 weeks vs 1x/week | Not statistically significant:<br>2.41±0.44 vs 2.5±0.42<br>MD: -0.09, 95%CI<br>[-0.36;0.18] (p=0.51)* | 1, 21 vs 19 § | Grgicevic, 1980 |
| | 3x/2 weeks vs < 1x/week | After 1 session (1 vs 1):<br>Not statistically significant:<br>2.64±0.56 vs 2.64±0.50 λ<br>MD: 0.0, 95%CI<br>[-0.24;0.24] (p=1.00)* | 1, 38 vs 38 § | Grgicevic, 1983 |
| | | After 7 sessions (7 vs 7):<br>Not statistically significant: | 1, 11 vs 12 § | |

| | | | | |
|---|---|---|---|---|
| | | 2.53±0.34 vs 2.46±0.36 λ<br>MD: 0.07, 95%CI<br>[-0.21;0.36] (p=0.61)* | | |
| | | After 15 sessions (15 vs 13):<br>Not statistically significant:<br>2.74±0.60 vs 2.86±0.60 λ<br>MD: -0.12, 95%CI<br>[-0.49;0.25] (p=0.53)* | 1, 28 vs 16 § | |
| | | After 35 sessions (36 vs 36):<br>Not statistically significant:<br>2.68±0.68 vs 2.67±0.58 λ<br>MD: 0.01, 95%CI<br>[-0.40;0.42] (p=0.96)* | 1, 16 vs 23 § | |
| | | After 75 sessions (70 vs 71):<br>Not statistically significant:<br>2.57±0.45 vs 2.43±0.46 λ<br>MD: 0.14, 95%CI<br>[-0.10;0.38] (p=0.26)* | 1, 25 vs 29 § | |
| | | After >100 sessions (175 vs 153):<br>Not statistically significant:<br>2.41±0.43 vs 2.50±0.41 λ<br>MD: -0.08, 95%CI<br>[-0.34;0.18] (p=0.53)* | 1, 21 vs 19 § | |
| Factor V [u/l x $10^3$] | 3x/2 weeks vs < 1x/week | After 1 session (1 vs 1):<br>Not statistically significant:<br>0.86±0.10 vs 0.86±0.10 λ<br>MD: 0.00, 95%CI<br>[-0.05;0.05] (p=0.95)* | 1, 38 vs 38 § | Grgicevic, 1983 |
| | | After 7 sessions (7 vs 7):<br><u>Statistically significant:</u><br>0.87±0.07 vs 0.93±0.07 λ<br>MD: -0.06, 95%CI<br>[-0.12;-0.00] (p=0.04)*<br>*In favour of plasma donation <1x/week* | 1, 11 vs 12 § | |
| | | After 15 sessions (15 vs 13):<br><u>Statistically significant:</u><br>0.80±0.01 vs 0.88±0.11 λ<br>MD: -0.07, 95%CI<br>[-0.14;-0.01] (p=0.03)* | 1, 22 vs 16 § | |

| | | | | |
|---|---|---|---|---|
| | | *In favour of plasma donation <1x/week* | | |
| | | After 35 sessions (36 vs 36):<br>Not statistically significant:<br>0.84±0.08 vs 0.89±0.10<br>MD: -0.05, 95%CI<br>[-0.11;0.01] (p=0.10)* | 1, 16 vs 23 § | |
| | | After 75 sessions (70 vs 71):<br>Statistically significant:<br>0.91±0.11 vs 0.84±0.11 λ<br>MD: 0.07, 95%CI<br>[0.01;0.13] (p=0.03)*<br>*In favour of plasma donation <3x/2 weeks* | 1, 24 vs 29 § | |
| | | After >100 sessions (175 vs 153):<br>Not statistically significant:<br>0.89±0.13 vs 0.82±0.14 λ<br>MD: 0.07, 95%CI<br>[-0.01;0.16] (p=0.08)* | 1, 21 vs 19 § | |
| Factor VIII [u/l x $10^3$] | 3x/2 weeks vs < 1x/week | After 1 session (1 vs 1):<br>Not statistically significant:<br>0.98±0.22 vs 0.97±0.22 λ<br>MD: 0.00, 95%CI<br>[-0.10;0.10] (p=0.96)* | 1, 37 vs 37 § | Grgicevic, 1983 |
| | | After 7 sessions (7 vs 7):<br>Not statistically significant:<br>0.91±0.18 vs 0.96±0.18 λ<br>MD: -0.05, 95%CI<br>[-0.20;0.10] (p=0.52)* | 1, 11 vs 12 § | |
| | | After 15 sessions (15 vs 13):<br>Not statistically significant:<br>0.98±0.18 vs 0.93±0.15 λ<br>MD: 0.05, 95%CI<br>[-0.05;0.16] (p=0.35)* | 1, 22 vs 16 § | |
| | | After 35 sessions (36 vs 36):<br>Not statistically significant:<br>1.05±0.21 vs 0.97±0.15 λ<br>MD: 0.08, 95%CI<br>[-0.04;0.21] (p=0.19)* | 1, 16 vs 22 § | |
| | | After 75 sessions (70 vs 71):<br>Not statistically significant: | 1, 24 vs 28 § | |

| | | | | |
|---|---|---|---|---|
| | | 0.95±0.13 vs 1.03±0.22 λ<br>MD: -0.08, 95%CI<br>[-0.17;0.02] (p=0.13)* | | |
| | | After >100 sessions (175 vs 153):<br>Not statistically significant:<br>0.95±0.16 vs 0.97±0.15 λ<br>MD: -0.02, 95%CI<br>[-0.12;0.07] (p=0.63)* | 1, 21 vs 19 § | |
| Alkaline phosphatase [u/l] | 3x/2 weeks vs 1x/week | Not statistically significant:<br>92.17±19.47 vs 85.4±15.99<br>MD: 6.77, 95%CI<br>[-3.29;16.83] (p=0.19)* | 1, 28 vs 20 § | Grgicevic, 1980 |
| GOT [u/l] | 3x/2 weeks vs 1x/week | Not statistically significant:<br>14.5±4.15 vs 14.68±6.79<br>MD: -0.18, 95%CI<br>[-3.54;3.18] (p=0.92)* | 1, 34 vs 19 § | Grgicevic, 1980 |
| GPT [u/l] | 3x/2 weeks vs 1x/week | Not statistically significant:<br>14.94±6.92 vs 15.06±7.76<br>MD: -0.12, 95%CI<br>[-4.39;4.15] (p=0.96)* | 1, 34 vs 18 § | Grgicevic, 1980 |

Mean ± SD, MD: mean difference, SD: standard deviation

* Calculations (MD, 95%CI) done by the reviewer(s) using Review Manager software

§ Imprecision (limited sample size)

λ data extracted from graph using WebPlotDigitizer

#### Twice per week versus no plasmapheresis for more than two years, for one to two years, or for one year or less

| Outcome | Comparison (Intervention vs comparator) | Effect size | #studies, #participants | Reference |
|---|---|---|---|---|
| IgG [g/l[a]] | 2x/week for 1-2 years vs ≤1 year | Not statistically significant:<br>7.32±1.87 vs 7.67±1.55<br>MD: -0.35, 95%CI<br>[-1.33;0.63] (p=0.48)* | 1, 24 vs 23 § | Salvaggio, 1971 |
| | 2x/week for >2 years vs ≤1 year | Not statistically significant:<br>8.60±1.83 vs 7.67±1.55<br>MD: 0.93, 95%CI<br>[-0.05;1.91] (p=0.06)* | 1, 23 vs 23 § | |
| | 2x/week for >2 years vs 1-2 years | Statistically significant:<br>8.60±1.83 vs 7.32±1.87<br>MD: 1.28, 95%CI<br>[0.22;2.34] (p=0.02)*<br>*In favour of 1-2 years* | 1, 23 vs 24 § | |
| | 2x/week for >2 years vs no plasmapheresis | Statistically significant:<br>8.60±1.83 vs 11.80±2.92<br>MD: -3.20, 95%CI<br>[-4.53;-1.87] (p<0.00001)*<br>*In favour of no plasmapheresis* | 1, 23 vs 27 § | |
| | 2x/week for 1-2 years vs no plasmapheresis | Statistically significant:<br>7.32±1.87 vs 11.80±2.92<br>MD: -4.48, 95%CI<br>[-5.81;-3.15] (p<0.00001)*<br>*In favour of no plasmapheresis* | 1, 24 vs 27 § | |
| | 2x/week for ≤1 year vs no plasmapheresis | Statistically significant:<br>7.67±1.55 vs 11.80±2.92<br>MD: -4.13, 95%CI<br>[-5.40;-2.86] (p<0.00001)*<br>*In favour of no plasmapheresis* | 1, 23 vs 27 § | |
| Low IgG levels (i.e. level below lower limits of normal range for both groups without | 2x/week for >2 years vs 1-2 years | Not statistically significant:<br>11/23 vs 16/24 §<br>RR: 0.72, 95%CI [0.43;1.20] ¥<br>(p=0.20) | 1, 23 vs 24 | Salvaggio, 1971 |

| | | | | |
|---|---|---|---|---|
| plasmapheresis (normal Louisiane prisoners) and Behring values (healthy European civilians) | | | | |
| | 2x/week for >2 years vs ≤1 year | Not statistically significant:<br>11/23 vs 15/23 §<br>RR: 0.73, 95%CI [0.44;1.23] ¥<br>(p=0.24) | 1, 23 vs 23 | |
| | 2x/week for 1-2 years vs ≤1 year | Not statistically significant:<br>16/24 vs 15/23 §<br>RR: 1.02, 95%CI [0.68;1.54] ¥<br>(p=0.92) | 1, 24 vs 23 | |
| | 2x/week for >2 years vs no plasmapheresis | <u>Statistically significant:</u><br>11/23 vs 2/25 §<br>RR: 5.98, 95%CI [1.48;24.15]<br>(p=0.01)<br>*In favour of no plasmapheresis* | 1, 23 vs 25 | |
| | 2x/week for 1-2 years vs no plasmapheresis | <u>Statistically significant:</u><br>16/24 vs 2/25 §<br>RR: 8.33, 95%CI [2.14;32.44]<br>(p=0.002)<br>*In favour of no plasmapheresis* | 1, 24 vs 25 | |
| | 2x/week for ≤1 year vs no plasmapheresis | <u>Statistically significant:</u><br>15/23 vs 2/25 §<br>RR: 8.15, 95%CI [2.09;31.84]<br>(p=0.003)<br>*In favour of no plasmapheresis* | 1, 23 vs 25 | |
| IgA [g/l[a]] | 2x/week for 1-2 years vs ≤1 year | Not statistically significant:<br>1.20±0.32 vs 1.28±0.43<br>MD: -0.08, 95%CI<br>[-0.30;0.14] (p=0.47)* | 1, 24 vs 23 § | Salvaggio, 1971 |
| | 2x/week for >2 years vs ≤1 year | Not statistically significant:<br>1.32±0.27 vs 1.28±0.43<br>MD: 0.04, 95%CI<br>[-0.17;0.25] (p=0.71)* | 1, 23 vs 23 § | |
| | 2x/week for >2 years vs 1-2 years | Not statistically significant:<br>1.32±0.27vs 1.20±0.32<br>MD: 0.12, 95%CI<br>[-0.05;0.29] (p=0.16)* | 1, 23 vs 24 § | |

| | | | | |
|---|---|---|---|---|
| | 2x/week for >2 years vs no plasmapheresis | Statistically significant:<br>1.32±0.27 vs 1.78±0.89<br>MD: -0.46, 95%CI<br>[-0.81;-0.11] (p=0.01)*<br>*In favour of no plasmapheresis* | 1, 23 vs 27 § | |
| | 2x/week for 1-2 years vs no plasmapheresis | Statistically significant:<br>1.20±0.32 vs 1.78±0.89<br>MD: -0.58, 95%CI<br>[-0.94;-0.22] (p=0.002)*<br>*In favour of no plasmapheresis* | 1, 24 vs 27 § | |
| | 2x/week for ≤1 year vs no plasmapheresis | Statistically significant:<br>1.28±0.43 vs 1.78±0.89<br>MD: -0.50, 95%CI<br>[-0.88;-0.12] (p=0.010)*<br>*In favour of no plasmapheresis* | 1, 23 vs 27 § | |
| Low IgA levels (i.e. level below lower limits of normal range for both groups without plasmapheresis (normal Louisiane prisoners) and Behring values (healthy European civilians) | 2x/week for >2 years vs 1-2 years | Not statistically significant:<br>10/23 vs 14/24 §<br>RR: 0.75, 95%CI [0.42;1.33] ¥<br>(p=0.32) | 1, 23 vs 24 | Salvaggio, 1971 |
| | 2x/week for >2 years vs ≤1 year | Not statistically significant:<br>10/23 vs 13/23 §<br>RR: 0.77, 95%CI [0.43;1.38] ¥<br>(p=0.38) | 1, 23 vs 23 | |
| | 2x/week for 1-2 years vs ≤1 year | Not statistically significant:<br>14/24 vs 13/23 §<br>RR: 1.03, 95%CI [0.63;1.69] ¥<br>(p=0.90) | 1, 24 vs 23 | |
| | 2x/week for >2 years vs no plasmapheresis | Not statistically significant:<br>10/23 vs 5/25 §<br>RR: 2.17, 95%CI [0.87;5.41] ¥<br>(p=0.10) | 1, 23 vs 25 | |
| | 2x/week for 1-2 years vs no plasmapheresis | Statistically significant:<br>14/24 vs 5/25 §<br>RR: 2.92, 95%CI [1.24;6.85]<br>(p=0.01) | 1, 24 vs 25 | |

| | | | | |
|---|---|---|---|---|
| | | *In favour of no plasmapheresis* | | |
| | 2x/week for ≤1 year vs no plasmapheresis | Statistically significant:<br>13/23 vs 5/25 §<br>RR: 2.83, 95%CI [1.19;6.69] (p=0.02)<br>*In favour of no plasmapheresis* | 1, 23 vs 25 | |
| IgM [g/l[a]] | 2x/week for 1-2 years vs ≤1 year | Not statistically significant:<br>0.88±0.46 vs 0.71±0.18<br>MD: 0.17, 95%CI [-0.03;0.37] (p=0.09)* | 1, 24 vs 23 § | Salvaggio, 1971 |
| | 2x/week for >2 years vs ≤1 year | Not statistically significant:<br>0.75±0.33 vs 0.71±0.18<br>MD: 0.04, 95%CI [-0.11;0.19] (p=0.61)* | 1, 23 vs 23 § | |
| | 2x/week for >2 years vs 1-2 years | Not statistically significant:<br>0.75±0.33 vs 0.88±0.46<br>MD: -0.13, 95%CI [-0.36;0.10] (p=0.26)* | 1, 23 vs 24 § | |
| | 2x/week for >2 years vs no plasmapheresis | Statistically significant:<br>0.75±0.33 vs 1.20±0.59<br>MD: -0.45, 95%CI [-0.71;-0.19] (p=0.0007)*<br>*In favour of no plasmapheresis* | 1, 23 vs 27 § | |
| | 2x/week for 1-2 years vs no plasmapheresis | Statistically significant:<br>0.88±46 vs 1.20±0.59<br>MD: -0.32, 95%CI [-0.61;-0.03] (p=0.03)*<br>*In favour of no plasmapheresis* | 1, 24 vs 27 § | |
| | 2x/week for ≤1 year vs no plasmapheresis | Statistically significant:<br>0.71±0.18 vs 1.20±0.59<br>MD: -0.49, 95%CI [-0.72;-0.26] (p<0.0001)*<br>*In favour of no plasmapheresis* | 1, 23 vs 27 § | |
| Low IgM levels (i.e. level below lower limits of normal range for both groups without plasmapheresis (normal Louisiane | 2x/week for >2 years vs 1-2 years | Not statistically significant:<br>17/23 vs 14/24 §<br>RR: 1.27, 95%CI [0.84;1.92] ¥ (p=0.27) | 1, 23 vs 24 | Salvaggio, 1971 |

| | | | | |
|---|---|---|---|---|
| prisoners) and Behring values (healthy European civilians) | | | | |
| | 2x/week for >2 years vs ≤1 year | Not statistically significant:<br>17/23 vs 17/23 §<br>RR: 1.00, 95%CI [0.71;1.41] ¥<br>(p=1.00) | 1, 23 vs 23 | |
| | 2x/week for 1-2 years vs ≤1 year | Not statistically significant:<br>14/24 vs 17/23 §<br>RR: 0.49, 95%CI [0.14;1.70] ¥<br>(p=0.26) | 1, 24 vs 23 | |
| | 2x/week for >2 years vs no plasmapheresis | Statistically significant:<br>17/23 vs 6/25 §<br>RR: 3.08, 95%CI [1.47;6.45]<br>(p=0.003)<br>*In favour of no plasmapheresis* | 1, 23 vs 25 | |
| | 2x/week for 1-2 years vs no plasmapheresis | Statistically significant:<br>14/24 vs 6/25 §<br>RR: 2.43, 95%CI [1.12;5.28]<br>(p=0.02)<br>*In favour of no plasmapheresis* | 1, 24 vs 25 | |
| | 2x/week for ≤1 year vs no plasmapheresis | Statistically significant:<br>17/23 vs 6/25 §<br>RR: 3.08, 95%CI [1.47;6.45]<br>(p=0.003)<br>*In favour of no plasmapheresis* | 1, 23 vs 25 | |
| Albumin [g/l[a]] | 2x/week for 1-2 years vs ≤1 year | Not statistically significant:<br>25.74±3.80 vs 27.73±3.23<br>MD: -1.99, 95%CI<br>[-4.00;0.02] (p=0.05)* | 1, 24 vs 23 § | Salvaggio, 1971 |
| | 2x/week for >2 years vs ≤1 year | Not statistically significant:<br>26.17±2.32 vs 27.73±3.23<br>MD: -1.56, 95%CI<br>[-3.19;0.07] (p=0.06)* | 1, 23 vs 23 § | |
| | 2x/week for >2 years vs 1-2 years | Not statistically significant:<br>26.17±2.32 vs 25.74±3.80<br>MD: 0.43, 95%CI<br>[-1.36;2.22] (p=0.64)* | 1, 23 vs 24 § | |

| | | | | |
|---|---|---|---|---|
| | 2x/week for >2 years vs no plasmapheresis | Statistically significant:<br>26.17±2.32 vs 31.99±4.57<br>MD: -5.82, 95%CI<br>[-7.79;-3.85] (p<0.00001)*<br>*In favour of no plasmapheresis* | 1, 23 vs 27 § | |
| | 2x/week for 1-2 years vs no plasmapheresis | Statistically significant:<br>25.74±3.80 vs 31.99±4.57<br>MD: -6.25, 95%CI<br>[-8.55;-3.95] (p<0.00001)*<br>*In favour of no plasmapheresis* | 1, 24 vs 27 § | |
| | 2x/week for ≤1 year vs no plasmapheresis | Statistically significant:<br>27.73±3.23 vs 31.99±4.57<br>MD: -4.26, 95%CI<br>[-6.43;-2.09] (p=0.0001)*<br>*In favour of no plasmapheresis* | 1, 23 vs 27 § | |
| B1-globulin [mg%[a]] | 2x/week for 1-2 years vs ≤1 year | Not statistically significant:<br>0.64±0.13 vs 0.70±0.14<br>MD: -0.06, 95%CI<br>[-0.14;0.02] (p=0.13)* | 1, 24 vs 23 § | Salvaggio, 1971 |
| | 2x/week for >2 years vs ≤1 year | Not statistically significant:<br>0.65±0.16 vs 0.70±0.14<br>MD: -0.05, 95%CI<br>[-0.14;0.03] (p=0.26)* | 1, 23 vs 23 § | |
| | 2x/week for >2 years vs 1-2 years | Not statistically significant:<br>0.65±0.16 vs 0.64±0.13<br>MD: 0.01, 95%CI<br>[-0.07;0.09] (p=0.81)* | 1, 23 vs 24 § | |
| | 2x/week for >2 years vs no plasmapheresis | Statistically significant:<br>0.65±0.16 vs 0.83±0.16<br>MD: -0.18, 95%CI<br>[-0.27;-0.09] (p<0.0001)*<br>*In favour of no plasmapheresis* | 1, 23 vs 27 § | |
| | 2x/week for 1-2 years vs no plasmapheresis | Statistically significant:<br>0.64±0.13 vs 0.83±0.16<br>MD: -0.19, 95%CI<br>[-0.27;-0.11] (p<0.00001)*<br>*In favour of no plasmapheresis* | 1, 24 vs 27 § | |

| | | | | |
|---|---|---|---|---|
| | 2x/week for ≤1 year vs no plasmapheresis | Statistically significant:<br>0.70±0.14 vs 0.83±0.16<br>MD: -0.13, 95%CI<br>[-0.21;-0.05] (p=0.002)*<br>*In favour of no plasmapheresis* | 1, 23 vs 27 § | |
| Transferrin [mg/dl] | 2x/week for 1-2 years vs ≤1 year | Not statistically significant:<br>178±23 vs 170±21<br>MD: 8, 95%CI<br>[-4.58;20.58] (p=0.21)* | 1, 24 vs 23 § | Salvaggio, 1971 |
| | 2x/week for >2 years vs ≤1 year | Not statistically significant:<br>173±38 vs 170±21<br>MD: 3, 95%CI<br>[-14.74;20.74] (p=0.74)* | 1, 23 vs 23 § | |
| | 2x/week for >2 years vs 1-2 years | Not statistically significant:<br>173±38 vs 178±23<br>MD: -5, 95%CI<br>[-23.05;13.05] (p=0.59)* | 1, 23 vs 24 § | |
| | 2x/week for >2 years vs no plasmapheresis | Not statistically significant:<br>173±38 vs 193±46<br>MD: -20, 95%CI<br>[-43.29;-3.29] (p=0.09)* | 1, 23 vs 27 § | |
| | 2x/week for 1-2 years vs no plasmapheresis | Not statistically significant:<br>178±23 vs 193±46<br>MD: -15, 95%CI<br>[-34.64;4.64] (p=0.13)* | 1, 24 vs 27 § | |
| | 2x/week for ≤1 year vs no plasmapheresis | Statistically significant:<br>170±21 vs 193±46<br>MD: -23, 95%CI<br>[-42.36;-3.64] (p=0.02)*<br>*In favour of no plasmapheresis* | 1, 23 vs 27 § | |
| Ceruloplasmin [mg/dl] | 2x/week for 1-2 years vs ≤1 year | Not statistically significant:<br>34±12 vs 31±8<br>MD: 3, 95%CI<br>[-2.81;8.81] (p=0.31)* | 1, 24 vs 23 § | Salvaggio, 1971 |
| | 2x/week for >2 years vs ≤1 year | Not statistically significant:<br>31±13 vs 31±8<br>MD: 0, 95%CI<br>[-6.24;6.24] (p=1)* | 1, 23 vs 23 § | |

| | | | | |
|---|---|---|---|---|
| | 2x/week for >2 years vs 1-2 years | Not statistically significant: 31±13 vs 34±12 MD: -3, 95%CI [-10.16;4.16] (p=0.41)* | 1, 23 vs 24 § | |
| | 2x/week for >2 years vs no plasmapheresis | Not statistically significant: 31±13 vs 31±11 MD: 0, 95%CI [-6.74;6.74] (p=1.00)* | 1, 23 vs 27 § | |
| | 2x/week for 1-2 years vs no plasmapheresis | Not statistically significant: 34±12 vs 31±11 MD: 3, 95%CI [-3.35;9.35] (p=0.35)* | 1, 24 vs 27 § | |
| | 2x/week for ≤1 year vs no plasmapheresis | Not statistically significant: 31±8 vs 31±11 MD: 0, 95%CI [-5.28;5.28] (p=1.00)* | 1, 23 vs 27 § | |
| Haptoglobin [mg/dl] | 2x/week for 1-2 years vs ≤1 year | Not statistically significant: 154±65 vs 145±48 MD: 9, 95%CI [-23.57;41.57] (p=0.59)* | 1, 24 vs 23 § | Salvaggio, 1971 |
| | 2x/week for >2 years vs ≤1 year | Not statistically significant: 137±41 vs 145±48 MD: -8, 95%CI [-33.8;17.8] (p=0.54)* | 1, 23 vs 23 § | |
| | 2x/week for >2 years vs 1-2 years | Not statistically significant: 137±41 vs 154±65 MD: -17, 95%CI [-47.94;13.94] (p=0.28)* | 1, 23 vs 24 § | |
| | 2x/week for >2 years vs no plasmapheresis | Not statistically significant: 137±41 vs 129±52 MD: 8, 95%CI [-17.80;33.80] (p=0.54)* | 1, 23 vs 27 § | |
| | 2x/week for 1-2 years vs no plasmapheresis | Not statistically significant: 154±65vs 129±52 MD: 25, 95%CI [-7.57;57.57] (p=0.13)* | 1, 24 vs 27 § | |
| | 2x/week for ≤1 year vs no plasmapheresis | Not statistically significant: 145±48 vs 129±52 MD: 16, 95%CI | 1, 23 vs 27 § | |

| | | |
|---|---|---|
| | | [-11.74;43.74] (p=0.26)* |

Mean ± SD, MD: mean difference, SD: standard deviation

* Calculations (MD, 95%CI) done by the reviewer(s) using Review Manager software

§ Imprecision (limited sample size or low number of events)

[a]Immunoglobulin and B1-globulin concentrations were expressed in mg% in the paper by Salvaggio et al. (1971). To facilitate comparison between study data, we converted mg% to g/l.

#### Plasma donation interval 2-4 days versus 5-9 days versus ≥10 days

| Outcome | Comparison (Intervention vs comparator) | Effect size | #studies, #participants | Reference |
|---|---|---|---|---|
| Predicted change in total cholesterol | 2-4 days vs 5-9 days in between donations (during 16 weeks) | Males:<br>High at baseline (≥240):<br>Statistically significant:<br>-32.1±4 vs -13.8±3.9 λ<br>MD: -18.3, 95%CI<br>[-22.37;-14.23] (p<0.00001)*<br>*In favour of 2-4 days in between donations* | 1, 9 vs 6 § | Rosa-Bray, 2013 |
| | | Males:<br>Higher than desired at baseline (200-239):<br>Statistically significant:<br>-23.5±2.1 vs -9±2 λ<br>MD: -14.5, 95%CI<br>[-15.7;-13.3] (p<0.00001)*<br>*In favour of 2-4 days in between donations* | 1, 27 vs 19 § | |
| | | Males:<br>Acceptable at baseline (<200): More<br>Statistically significant:<br>-5±1.1 vs 6.9±1.5 λ<br>MD: -11.9, 95%CI<br>[-12.29;-11.51] (p<0.00001)*<br>*In favour of 2-4 days in between donations* | 1, 110 vs 77 § | |
| | | Females:<br>High at baseline (≥240):<br>Statistically significant:<br>-46.6±5.2 vs -32±5 λ<br>MD: -14.6, 95%CI<br>[-22.26;-6.94] (p=0.0002)*<br>*In favour of 2-4 days in between donations* | 1, 3 vs 4 § | |
| | | Females:<br>Higher than desired at baseline (200-239):<br>Statistically significant:<br>-22±2.8 vs -10.7±2.7 λ<br>MD: -11.3, 95%CI | 1, 12 vs 15 § | |

| | | | |
|---|---|---|---|
| | | [-13.39;-9.21] (p<0.00001)*<br>*In favour of 2-4 days in between donations* | |
| | | Females:<br>Acceptable at baseline (<200):<br><u>Statistically significant:</u><br>-7.3±1.6 vs 1.6±1.4 λ<br>MD: -8.9, 95%CI<br>[-9.54;-8.26] (p<0.00001)*<br>*In favour of 2-4 days in between donations* | 1, 39 vs 48 § |
| | 2-4 days vs ≥ 10 days in between donations (during 16 weeks) | Males:<br>High at baseline (≥240):<br><u>Statistically significant:</u><br>-32.1±4 vs -17±3.8 λ<br>MD: -15.1, 95%CI<br>[-18.7;-11.5] (p<0.00001)*<br>*In favour of 2-4 days in between donations* | 1, 9 vs 9 § |
| | | Males:<br>Higher than desired at baseline (200-239):<br><u>Statistically significant:</u><br>-23.5±2.1 vs -2.8±2.2 λ<br>MD: -20.7, 95%CI<br>[-21.83;-19.57] (p<0.00001)*<br>*In favour of 2-4 days in between donations* | 1, 27 vs 29 § |
| | | Males:<br>Acceptable at baseline (<200):<br><u>Statistically significant:</u><br>-5±1.1 vs 7.8±1 λ<br>MD: -12.8, 95%CI<br>[-13.07;-12.53] (p<0.00001)*<br>*In favour of 2-4 days in between donations* | 1, 110 vs 121 § |
| | | Females:<br>High at baseline (≥240):<br><u>Statistically significant:</u><br>-46.6±5.2 vs -20.8±5.1 λ<br>MD: -25.8, 95%CI<br>[-32.79;-18.81] (p<0.00001)*<br>*In favour of 2-4 days in between donations* | 1, 3 vs 7 § |
| | | Females:<br>Higher than desired at baseline (200-239): | 1, 12 vs 30 § |

| | | | |
|---|---|---|---|
| | | Statistically significant:<br>-22±2.8 vs -7.3±2.6 λ<br>MD: -14.7, 95%CI<br>[-16.54;-12.86] (p<0.00001)*<br>*In favour of 2-4 days in between donations* | |
| | | Females:<br>Acceptable at baseline (<200):<br>Statistically significant:<br>-7.3±1.6 vs 3.3±1.7 λ<br>MD: -10.6, 95%CI<br>[-11.2;-10] (p<0.00001)*<br>*In favour of 2-4 days in between donations* | 1, 39 vs 98 § |
| | 5-9 days vs ≥ 10 days in between donations (during 16 weeks) | Males:<br>High at baseline (≥240):<br>Not statistically significant:<br>-13.8±3.9 vs -17±3.8 λ<br>MD: 3.2, 95%CI<br>[-0.79;7.19] (p=0.12)* | 1, 6 vs 9 § |
| | | Males:<br>Higher than desired at baseline (200-239):<br>Statistically significant:<br>-9±2 vs -2.8±2.2 λ<br>MD: -6.2, 95%CI<br>[-7.4;-5] (p<0.00001)*<br>*In favour of 5-9 days in between donations* | 1, 19 vs 29 § |
| | | Males:<br>Acceptable at baseline (<200):<br>Statistically significant:<br>6.9±1.5 vs 7.8±1 λ<br>MD: -0.9, 95%CI<br>[-1.28;-0.52] (p<0.00001)*<br>*In favour of 5-9 days in between donations* | 1, 77 vs 121 § |
| | | Females:<br>High at baseline (≥240):<br>Statistically significant:<br>-32±5 vs -20.8±5.1 λ<br>MD: -11.2, 95%CI<br>[-17.39;-5.01] (p=0.0004)*<br>*In favour of 2-4 days in between donations* | 1, 5 vs 7 § |

| | | | | |
|---|---|---|---|---|
| | | Females:<br>Higher than desired at baseline (200-239):<br><u>Statistically significant:</u><br>-10.7±2.7 vs -7.3±2.6 λ<br>MD: -3.4, 95%CI<br>[-5.05;-1.75] ($p<0.0001$)*<br>*In favour of 2-4 days in between donations* | 1, 15 vs 30 § | |
| | | Females:<br>Acceptable at baseline (<200):<br><u>Statistically significant:</u><br>1.6±1.4 vs 3.3±1.7 λ<br>MD: -1.7, 95%CI<br>[-2.22;-1.18] ($p<0.00001$)*<br>*In favour of 2-4 days in between donations* | 1, 48 vs 98 § | |
| Predicted change in LDL cholesterol | 2-4 days vs 5-9 days in between donations (during 16 weeks) | Males:<br>High at baseline (≥160):<br><u>Statistically significant:</u><br>-34.6±3.8 vs -17.8±4 λ<br>MD: -16.8, 95%CI<br>[-20.85;-12.75] ($p<0.00001$)*<br>*In favour of 2-4 days in between donations* | 1, 9 vs 6 § | Rosa-Bray, 2013 |
| | | Males:<br>Higher than desired at baseline (130-159):<br><u>Statistically significant:</u><br>-15.3±1.9 vs -4.6±1.9 λ<br>MD: -10.7, 95%CI<br>[-11.88;-9.52] ($p<0.00001$)*<br>*In favour of 2-4 days in between donations* | 1, 24 vs 17 § | |
| | | Males:<br>Acceptable at baseline (<130):<br><u>Statistically significant:</u><br>-2.2±1 vs 5.4±1 λ<br>MD: -7.6, 95%CI<br>[-7.89;-7.31] ($p<0.00001$)*<br>*In favour of 2-4 days in between donations* | 1, 112 vs 79 § | |
| | | Females:<br>High at baseline (≥160):<br><u>Statistically significant:</u><br>-35.3±5 vs -20.8±4.8 λ | 1, 3 vs 4 § | |

| | | | |
|---|---|---|---|
| | | MD: -14.5, 95%CI<br>[-21.86;-7.14] (p=0.0001)*<br>*In favour of 2-4 days in between donations* | |
| | | Females:<br>Higher than desired at baseline (130-159):<br>Statistically significant:<br>-18.4±2.8 vs -10.5±2.5 λ<br>MD: -7.9, 95%CI<br>[-10.25;-5.55] (p<0.00001)*<br>*In favour of 2-4 days in between donations* | 1, 9 vs 11 § |
| | | Females:<br>Acceptable at baseline (<130):<br>Statistically significant:<br>-1.9±0.6 vs 3.7±0.9 λ<br>MD: -5.6, 95%CI<br>[-5.91;-5.29] (p<0.00001)*<br>*In favour of 2-4 days in between donations* | 1, 41 vs 51 § |
| | 2-4 days vs ≥ 10 days in between donations (during 16 weeks) | Males:<br>High at baseline (≥160):<br>Statistically significant:<br>-34.6±3.8 vs -19.1±4 λ<br>MD: -15.5, 95%CI<br>[-19.01;-11.99] (p<0.00001)*<br>*In favour of 2-4 days in between donations* | 1, 9 vs 10 § |
| | | Males:<br>Higher than desired at baseline (130-159):<br>Statistically significant:<br>-15.3±1.9 vs -0.4±1.8 λ<br>MD: -14.9, 95%CI<br>[-15.92;-13.88] (p<0.00001)*<br>*In favour of 2-4 days in between donations* | 1, 24 vs 27 § |
| | | Males:<br>Acceptable at baseline (<130):<br>Statistically significant:<br>-2.2±1 vs 6.8±1.2 λ<br>MD: -9, 95%CI<br>[-9.28;-8.72] (p<0.00001)*<br>*In favour of 2-4 days in between donations* | 1, 112 vs 124 § |

| | | | |
|---|---|---|---|
| | | Females:<br>High at baseline (≥160):<br><u>Statistically significant:</u><br>-35.3±5 vs -25.6±5 λ<br>MD: -9.7, 95%CI<br>[-16.33;-3.07] (p=0.004)*[a]<br>*In favour of 2-4 days in between donations* | 1, 5 vs 8 § |
| | | Females:<br>Higher than desired at baseline (130-159):<br><u>Statistically significant:</u><br>-18.4±2.8 vs -7.3±2.6 λ<br>MD: -11.1, 95%CI<br>[-13.22;-8.98] (p<0.00001)*<br>*In favour of 2-4 days in between donations* | 1, 9 vs 23 § |
| | | Females:<br>Acceptable at baseline (<130):<br><u>Statistically significant:</u><br>-1.9±0.6 vs 3±0.8 λ<br>MD: -4.9, 95%CI<br>[-5.14;-4.66] (p<0.00001)*<br>*In favour of 2-4 days in between donations* | 1, 41 vs 104 § |
| | 5-9 days vs ≥ 10 days in between donations (during 16 weeks) | Males:<br>High at baseline (≥160):<br>Not statistically significant:<br>-17.8±4 vs -19.1±4 λ<br>MD: 1.3, 95%CI<br>[-2.75;5.35] (p=0.53)* | 1, 6 vs 10 § |
| | | Males:<br>Higher than desired at baseline (130-159):<br><u>Statistically significant:</u><br>-4.6±1.9 vs -0.4±1.8 λ<br>MD: -4.2, 95%CI<br>[-5.33;-3.07] (p<0.00001)*<br>*In favour of 5-9 days in between donations* | 1, 17 vs 27 § |
| | | Males:<br>Acceptable at baseline (<130):<br><u>Statistically significant:</u><br>5.4±1 vs 6.8±1.2 λ<br>MD: -1.4, 95%CI | 1, 79 vs 124 § |

| | | | | |
|---|---|---|---|---|
| | | [-1.71;-1.09] (p<0.00001)*<br>*In favour of 5-9 days in between donations* | | |
| | | Females:<br>High at baseline (≥160):<br>Not statistically significant:<br>-20.8±4.8 vs -25.6±5 λ<br>MD: 4.8, 95%CI<br>[-1.04;10.64] (p=0.11)*<br>*In favour of 5-9 days in between donations* | 1, 4 vs 8 § | |
| | | Females:<br>Higher than desired at baseline (130-159):<br><u>Statistically significant:</u><br>-10.5±2.5 vs -7.3±2.6 λ<br>MD: -3.2, 95%CI<br>[-5.02;-1.38] (p=0.0006)*<br>*In favour of 5-9 days in between donations* | 1, 11 vs 12 § | |
| | | Females:<br>Acceptable at baseline (<130):<br><u>Statistically significant:</u><br>3.7±0.9 vs 3±0.8 λ<br>MD: 0.7, 95%CI<br>[0.41;0.99] (p<0.00001)*<br>*In favour of ≥10 days in between donations* | 1, 51 vs 104 § | |
| Predicted change in HDL cholesterol | 2-4 days vs 5-9 days in between donations (during 16 weeks) | Males:<br>Low at baseline (<40):<br><u>Statistically significant:</u><br>2.3±0.6 vs 4.7±0.8 λ<br>MD: -2.4, 95%CI<br>[-2.76;-2.04] (p<0.00001)*<br>*In favour of 5-9 days in between donations* | 1, 38 vs 27 § | Rosa-Bray, 2013 |
| | | Males:<br>Average at baseline (40-60):<br><u>Statistically significant:</u><br>-3.2±0.5 vs 0.3±0.6 λ<br>MD: -3.5, 95%CI<br>[-3.68;-3.32] (p<0.00001)*<br>*In favour of 5-9 days in between donations* | 1, 89 vs 63 § | |
| | | Males:<br>Optimal at baseline (>60): | 1, 17 vs 12 § | |

| | | | |
|---|---|---|---|
| | | <u>Statistically significant:</u><br>-9.7±1.4 vs -5.4±1.5 λ<br>MD: -4.3, 95%CI<br>[-5.38;-3.22] (p<0.00001)*<br>*In favour of 5-9 days in between donations* | |
| | | Females:<br>Low at baseline (<50):<br><u>Statistically significant:</u><br>0±0.7 vs 2.7±0.6 λ<br>MD: -2.7, 95%CI<br>[-4.83;-0.57] (p=0.01)*[a]<br>*In favour of 5-9 days in between donations* | 1, 25 vs 31 § |
| | | Females:<br>Average at baseline (50-60):<br><u>Statistically significant:</u><br>-4.1±0.7 vs -0.6±0.7 λ<br>MD: -3.5, 95%CI<br>[-3.93;-3.07] (p<0.00001)*<br>*In favour of 5-9 days in between donations* | 1, 19 vs 23 § |
| | | Females:<br>Optimal at baseline (>60):<br><u>Statistically significant:</u><br>-10.8±1.5 vs -6.7±1.5 λ<br>MD: -4.1, 95%CI<br>[-5.36;-2.84] (p<0.00001)*<br>*In favour of 5-9 days in between donations* | 1, 10 vs 12 § |
| | 2-4 days vs ≥ 10 days in between donations (during 16 weeks) | Males:<br>Low at baseline (<40):<br><u>Statistically significant:</u><br>2.3±0.6 vs 4.9±0.5 λ<br>MD: -2.6, 95%CI<br>[-2.84;-2.36] (p<0.00001)*<br>*In favour of ≥10 days in between donations* | 1, 38 vs 42 § |
| | | Males:<br>Average at baseline (40-60):<br><u>Statistically significant:</u><br>-3.2±0.5 vs 0.3±0.5 λ<br>MD: -3.5, 95%CI<br>[-3.64;-3.36] (p<0.00001)* | 1, 89 vs 99 § |

| | | | |
|---|---|---|---|
| | | *In favour of ≥10 days in between donations* | |
| | | Males:<br>Optimal at baseline (>60):<br><u>Statistically significant:</u><br>-9.7±1.4 vs -3.5±1.6 λ<br>MD: -6.2, 95%CI<br>[-7.18;-5.22] (p<0.00001)*<br>*In favour of ≥10 days in between donations* | 1, 17 vs 19 § |
| | | Females:<br>Low at baseline (<50):<br><u>Statistically significant:</u><br>0±0.7 vs 3.6±0.8 λ<br>MD: -3.6, 95%CI<br>[-3.94;-3.26] (p<0.00001)*<br>*In favour of ≥10 days in between donations* | 1, 25 vs 64 § |
| | | Females:<br>Average at baseline (50-60):<br><u>Statistically significant:</u><br>-4.1±0.7 vs 0.1±0.7 λ<br>MD: -4.2, 95%CI<br>[-4.57;-3.83] (p<0.00001)*<br>*In favour of ≥10 days in between donations* | 1, 19 vs 48 § |
| | | Females:<br>Optimal at baseline (>60):<br><u>Statistically significant:</u><br>-10.8±1.5 vs -4±1.5 λ<br>MD: -6.8, 95%CI<br>[-7.91;-5.69] (p<0.00001)*<br>*In favour of ≥10 days in between donations* | 1, 10 vs 24 § |
| | 5-9 days vs ≥ 10 days in between donations (during 16 weeks) | Males:<br>Low at baseline (<40):<br>Not statistically significant:<br>4.7±0.8 vs 4.9±0.5 λ<br>MD: -0.2, 95%CI<br>[-0.51;0.14] (p=0.25)* | 1, 27 vs 42 § |
| | | Males:<br>Average at baseline (40-60):<br>Not statistically significant:<br>0.3±0.6 vs 0.3±0.5 λ | 1, 63 vs 99 § |

| | | | |
|---|---|---|---|
| | | MD: 0, 95%CI<br>[-0.18;-0.18] (p=1)* | |
| | | Males:<br>Optimal at baseline (>60):<br><u>Statistically significant:</u><br>-5.4±1.5 vs -3.5±1.6 λ<br>MD: -1.9, 95%CI<br>[-3.01;-0.79] (p=0.0008)*<br>*In favour of ≥10 days in between donations* | 1, 12 vs 19 § |
| | | Females:<br>Low at baseline (<50):<br><u>Statistically significant:</u><br>2.7±0.6 vs 3.6±0.8 λ<br>MD: -0.9, 95%CI<br>[-1.19;-0.61] (p<0.00001)*<br>*In favour of ≥10 days in between donations* | 1, 31 vs 64 § |
| | | Females:<br>Average at baseline (50-60):<br><u>Statistically significant:</u><br>-0.6±0.7 vs 0.1±0.7 λ<br>MD: -0.7, 95%CI<br>[-1.05;-0.35] (p<0.0001)*<br>*In favour of ≥10 days in between donations* | 1, 23 vs 48 § |
| | | Females:<br>Optimal at baseline (>60):<br><u>Statistically significant:</u><br>-6.7±1.5 vs -4±1.5 λ<br>MD: -2.7, 95%CI<br>[-3.74;-1.66] (p<0.00001)*<br>*In favour of ≥10 days in between donations* | 1, 12 vs 24 § |

Mean ± SD, MD: mean difference, SD: standard deviation

* Calculations (MD, 95%CI) done by the reviewer(s) using Review Manager software

§ Imprecision (limited sample size)

λ data extracted from graph using WebPlotDigitizer

[a]Not statistically significant after applying the Holm-Bonferroni method. Given the multitude of pairwise comparisons that we performed using the data of Rosa-Bray (2013), the Holm-Bonferroni method was used to counteract the problem of multiple testing. Given that most p-values were very small, significance was unaffected by multiple testing corrections in all but two pairwise comparisons. The unadjusted p-values are shown.
